# Extremely potent monoclonal antibodies neutralize Omicron and other SARS-CoV-2 variants

**DOI:** 10.1101/2022.01.12.22269023

**Authors:** Zhaochun Chen, Peng Zhang, Yumiko Matsuoka, Yaroslav Tsybovsky, Kamille West, Celia Santos, Lisa F. Boyd, Hanh Nguyen, Anna Pomerenke, Tyler Stephens, Adam S. Olia, Valeria De Giorgi, Michael R. Holbrook, Robin Gross, Elena Postnikova, Nicole L. Garza, Reed F. Johnson, David H. Margulies, Peter D. Kwong, Harvey J. Alter, Ursula J. Buchholz, Paolo Lusso, Patrizia Farci

## Abstract

The ongoing coronavirus disease 2019 (COVID-19) pandemic, caused by severe acute respiratory syndrome coronavirus 2 (SARS-CoV-2), has triggered a devastating global health, social and economic crisis. The RNA nature and broad circulation of this virus facilitate the accumulation of mutations, leading to the continuous emergence of variants of concern with increased transmissibility or pathogenicity^1^. This poses a major challenge to the effectiveness of current vaccines and therapeutic antibodies^1, 2^. Thus, there is an urgent need for effective therapeutic and preventive measures with a broad spectrum of action, especially against variants with an unparalleled number of mutations such as the recently emerged Omicron variant, which is rapidly spreading across the globe^3^. Here, we used combinatorial antibody phage-display libraries from convalescent COVID-19 patients to generate monoclonal antibodies against the receptor-binding domain of the SARS-CoV-2 spike protein with ultrapotent neutralizing activity. One such antibody, NE12, neutralizes an early isolate, the WA-1 strain, as well as the Alpha and Delta variants with half-maximal inhibitory concentrations at picomolar level. A second antibody, NA8, has an unusual breadth of neutralization, with picomolar activity against both the Beta and Omicron variants. The prophylactic and therapeutic efficacy of NE12 and NA8 was confirmed in preclinical studies in the golden Syrian hamster model. Analysis by cryo-EM illustrated the structural basis for the neutralization properties of NE12 and NA8. Potent and broadly neutralizing antibodies against conserved regions of the SARS-CoV-2 spike protein may play a key role against future variants of concern that evade immune control.

---

The coronavirus disease 2019 (COVID-19) pandemic has caused more than 800,000 deaths in the United States and over 5 million worldwide^4^. Although effective vaccines against SARS-CoV-2 have been developed and deployed at an unprecedented pace, hesitancy in vaccination and a limited supply in developing countries have made the fight against this virus particularly challenging. In addition, like other RNA viruses, SARS-CoV-2 is subjected to continuous genetic variation with accumulation of mutations leading to immune evasion^1, 2^. As a result of viral evolution, the initial SARS-CoV-2 genetic lineages identified early during the pandemic in Wuhan^5^, China, have been progressively replaced by several variants of concern (VOCs), such as B.1.1.7 (Alpha), B.1.351 (Beta), P.1, (Gamma) and B.1.617.2 (Delta). The most recent VOC, first identified in South Africa and named Omicron (B.1.1.529), is rapidly spreading worldwide^3^. VOCs contain mutations that may increase transmissibility, induce resistance to vaccine-elicited antibodies and monoclonal antibodies (mAbs) and, potentially, enhance pathogenicity^6–8^. Of particular concern currently is the Omicron variant, which has an unprecedented number of mutations in the spike (S) protein^9^. Several recent studies have shown that the Omicron variant has reduced or abrogated sensitivity to neutralization by most mAbs, convalescent sera, and vaccine-elicited antibodies^10–15^. The resistance of emergent VOCs to current therapeutic mAbs and the reduced effectiveness of vaccines highlight the urgent need for the discovery of broadly neutralizing mAbs against SARS-CoV-2.

Most mAbs against SARS-CoV-2 have been isolated by using single B-cell cloning^2, 16^. We took advantage of our extensive experience in generating mAbs using combinatorial phage-display library technology^17, 18^ to generate mAbs from convalescent COVID-19 patients with high titers of neutralizing antibodies. Here, we report the isolation of mAbs that potently neutralize diverse and highly transmissible SARS-CoV-2 VOCs, including B.1.351 and B.1.1.529, and demonstrate both protective and therapeutic activity in a hamster preclinical model.

## Generation of anti-SARS-CoV-2 monoclonal antibodies using phage-display libraries from COVID-19 convalescent patients

Peripheral blood samples were collected from 12 convalescent COVID-19 patients with high neutralizing serum antibody titers against SARS-CoV-2^19^ (Extended Data Table 1). Total RNA was extracted from peripheral blood mononuclear cells (PBMC) and used for the construction of phage-display Fab libraries (Extended Data Fig. 1). A total of four phage-display Fab libraries, derived from either a single donor or multiple donors combined, were constructed, each consisting of 10^7^ to 10^9^ individual clones (Extended Data Fig. 2). Three sequential cycles of panning were carried out to enrich for specific clones using a stabilized trimeric spike protein (S- 2P) derived from the original SARS-CoV-2 strain, Wuhan-Hu-1 (GenBank accession number MN908947; S-protein sequence identical to that of the WA-1 strain)^20^. Subsequent screening of 672 individual clones by ELISA resulted in the identification of 538 clones that specifically bound to the S-2P protein. DNA sequencing identified 18 unique clones with distinct sequences. These clones were subcloned and expressed both as soluble Fabs and as complete IgG1 antibodies. The binding specificity of the cloned IgGs was confirmed by ELISA (Fig. 1a). The binding affinity of the 18 Fabs for the soluble S-2P trimer was further assessed by surface plasmon resonance (SPR). Six Fabs showed either poor or no binding by SPR while the remaining 12 exhibited high-affinity binding with equilibrium dissociation constants (*KD*) in the picomolar range for 10 and in the low nanomolar range for 2 (Extended Data Fig. 3 and Extended Data Table 2).

**Fig. 1.**
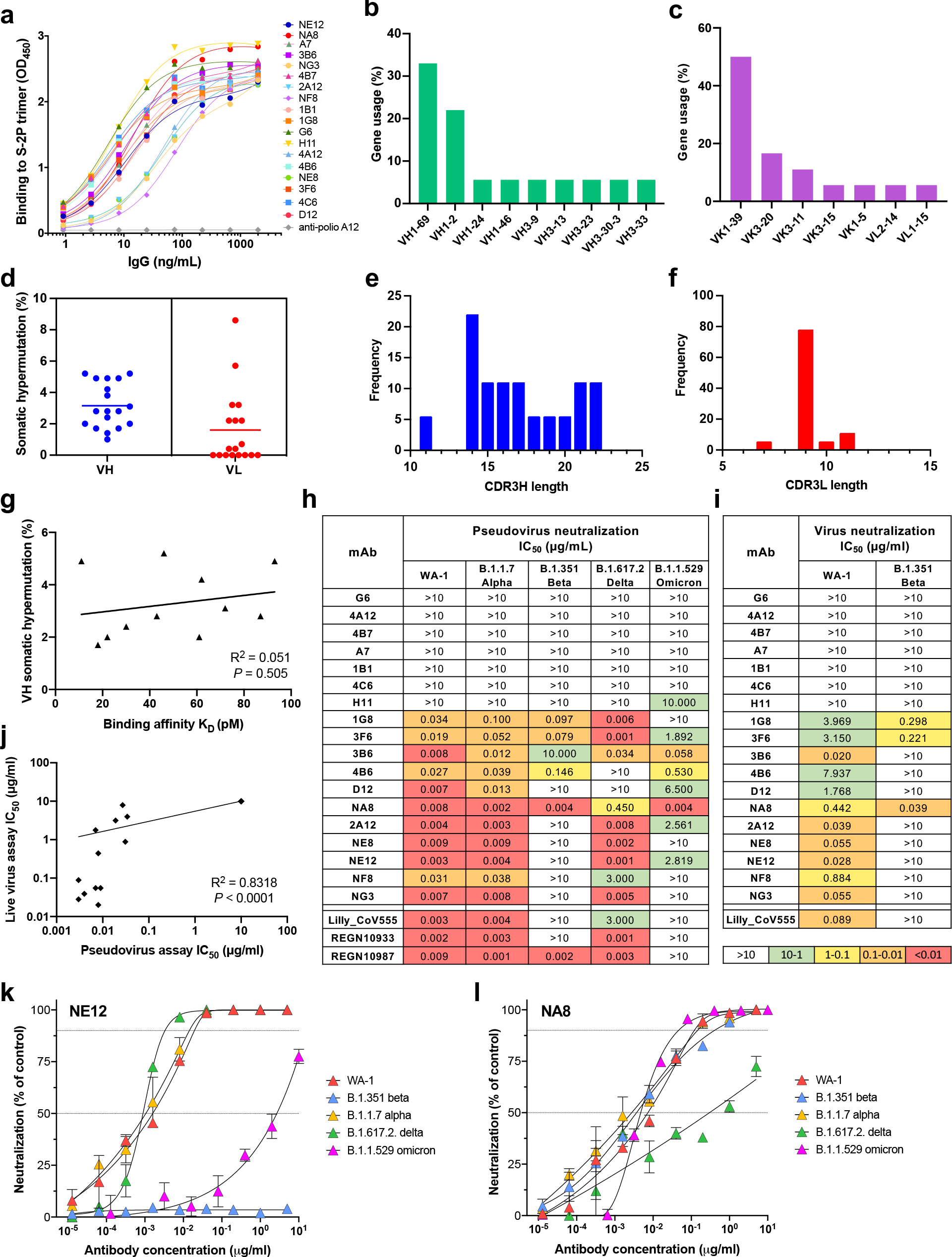
Genetic and functional characterization of mAbs against the SARS-CoV-2 spike protein. **a**, Normalized binding of 18 mAbs to a stabilized SARS-CoV-2 spike protein trimer, S- 2P, in ELISA. OD denotes optical density. **b,c,** V-gene usage of heavy (b) and light chains (c) of 18 mAbs against the SARS-CoV-2 spike protein. **d,** Rate of somatic hypermutation of V genes of heavy chains (VH) and light chains (VL) of 18 mAbs. **e,f,** Amino acid length of the CDR3 loop of heavy (CDR3H) (**e**) and light chain (CDR3L) (**f**) in 18 mAbs. **g,** Correlation between antibody binding affinity and frequency of VH gene somatic hypermutation. **h,** Heat map of pseudovirus neutralization activity. IC50 values (µg/mL) are shown for neutralization of SARS-CoV-2 pseudoviruses bearing spike proteins from the USA/WA1/2020 isolate (WA-1, lineage A), the Alpha variant (lineage B.1.1.7), Beta variant (B.1.351), Delta variant (B.1.617.2) and Omicron variant (B.1.1.529). The clinically approved neutralizing mAbs Lilly_CoV555, REGN10933 and REGN10987 were used in parallel for reference. **i,** Heat map of live virus neutralization against SARS-CoV-2 WA-1 and B.1.351. **j,** Correlation between neutralizing activities of mAbs in the pseudovirus and live virus assays. **k,l,** Neutralization curves of mAbs NE12 (**k**) and NA8 (**l**). The symbols indicate means from three independent experiments performed in duplicate. The dotted horizontal lines indicate the IC90 and IC50.

**Fig. 2.**
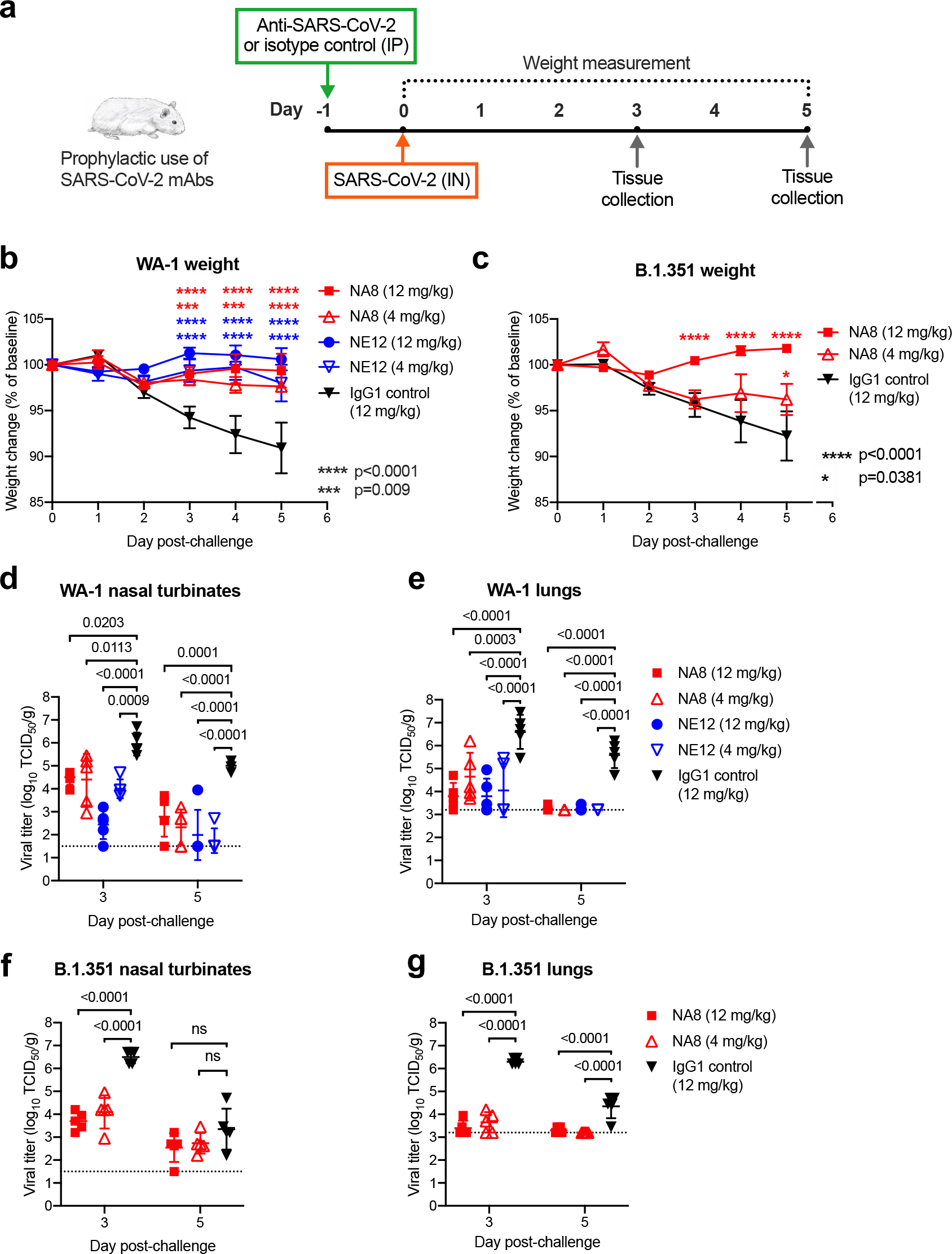
Prophylactic efficacy of two neutralizing mAbs, NE12 and NA8, against infection with SARS-CoV-2 in the golden Syrian hamster model. a, Design of the study. MAbs NE12 or NA8 were administered intraperitoneally (IP) at the dose of 12 or 4 mg/kg. Control animals received an anti-HIV-1 IgG1 (VRC01) at 12 mg/kg. One day later, each group of 10 animals was challenged with 10^4.5^ TCID50 of SARS-CoV-2 WA-1 or B.1.351, instilled intranasally (IN). The animal weight was monitored daily (days 0-3: n=10 per group; days 4, 5: n=5 per group) as an indicator of disease progression. Tissues were collected at 3 and 5 days after challenge for virus quantification (n=5 per group per time point). b,c, Body weight changes (means ± standard error) from baseline in hamsters that received neutralizing mAbs at different doses or isotype control (IgG) after challenge with SARS-CoV-2 WA-1 (b) or B.1.351 (c). d-g, Viral titers in nasal turbinate and lung tissues at day 3 and 5 post-infection with WA-1 (d,e) or the B.1.351 variant (f,g), as determined using an assay to quantify the TCID50 of infectious virus. Individual titers and means ± standard deviation (SD) are shown for each group. Dashed lines indicate the limit of detection of the assay. Statistical comparisons of weight changes between treatment groups were performed using a repeated-measures mixed-effects model with Tukey’s multiple- comparison test (panel b and c). Comparisons of the viral titers between groups were performed using two-way analysis of variance (ANOVA) with Tukey’s multiple-comparison test. P values between groups treated with neutralizing mAbs and isotype control are indicated. p>0.05, not significant (ns); * p=0.0381; *** P=0.0009; ****p<0.0001.

**Fig. 3.**
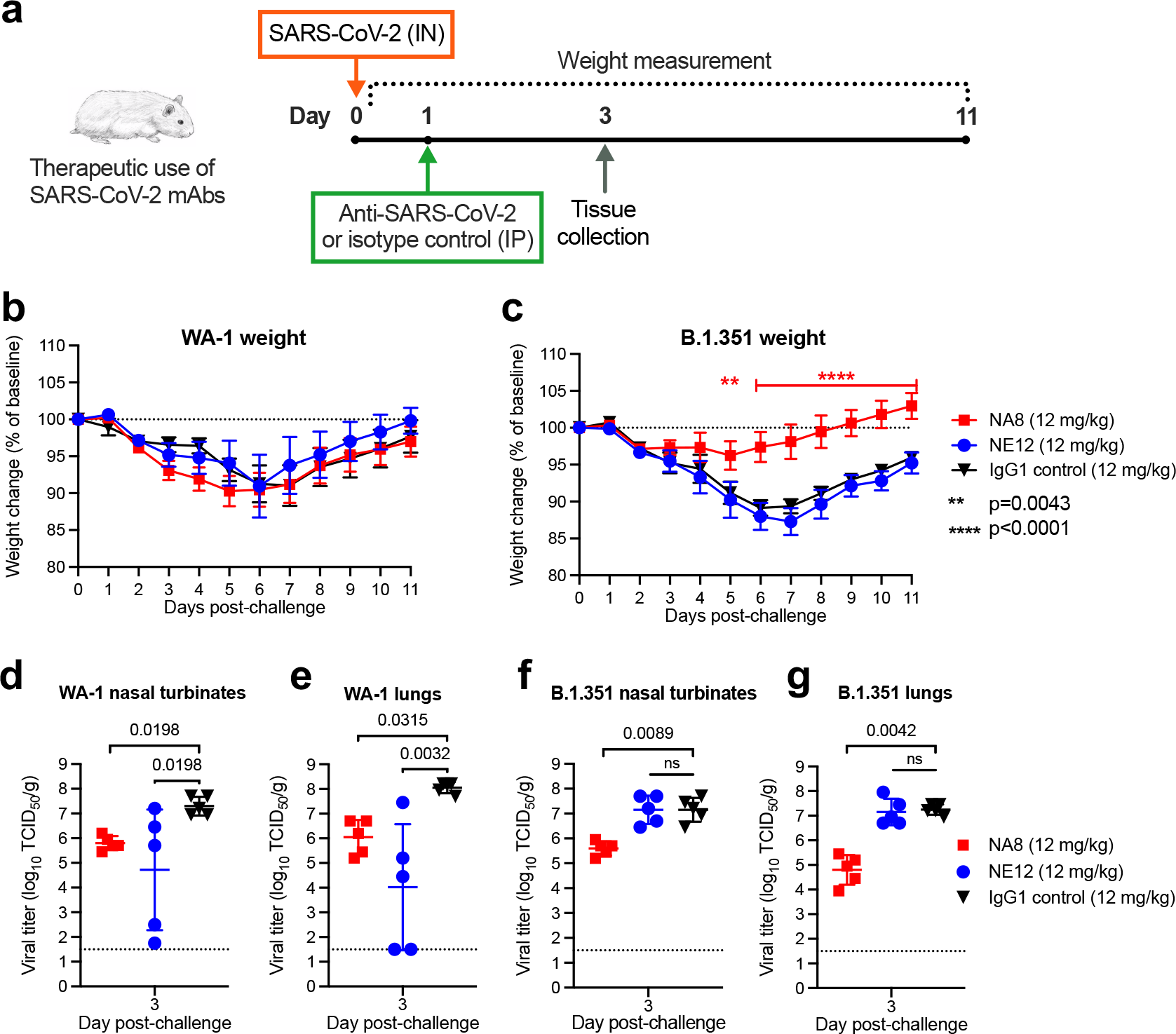
Therapeutic efficacy of two neutralizing mAbs, NE12 and NA8, against SARS-CoV- 2 in the golden Syrian hamster model. **a,** Design of the study. Each group of 10 animals was first challenged with 10^4.5^ TCID50 of SARS-CoV-2 WA-1 or B.1.351/Beta, instilled intranasally (IN). Twenty-four hours later, the animals were injected intraperitoneally (IP) with mAbs NE12 or NA8 at the dose of 12 mg/kg. Control animals received an anti-HIV-1 IgG1 (VRC01) at 12 mg/kg. The animal weight was monitored daily as an indicator of disease progression (days 0-2: n=10 per group; days 3-11: n=5 per group, except for the WA-1/NE12 group (days 6-11: n=3 due to a technical issue). Tissues were collected at day 3 after challenge for virus quantification (n=5 per group). **b,c,** Body weight changes (means ± standard error) from baseline in hamsters that were challenged with SARS-CoV-2 WA-1 (**b**) or B.1.351/Beta (**c**). **d-g,** Quantification of viral titers in nasal turbinate and lung tissues at day 3 post-challenge in animals infected with WA-1 (**d,e**) or the B.1.351/Beta variant (**f,g**), as determined using an assay to quantify the TCID50 of infectious virus. Individual titers and means ± SDs are shown for each group. Dashed lines indicate the limit of detection of the assay. Statistical comparisons of weight changes between treatment groups were performed using a repeated-measures mixed-effects model with Tukey’s multiple-comparison test. Comparisons of viral titers between the treatment groups were performed using the non-parametric Kruskal-Wallis test with a Benjamini, Krieger and Yekutieli false-discovery test. P values between groups treated with neutralizing mAbs and isotype control are indicated. p>0.05, not significant (ns); ** p=0.0043; **** p<0.0001

## Genetic characterization of anti-SARS-CoV-2 monoclonal antibodies

Genetic analysis of V genes of all 18 monoclonal antibodies indicates that the clones preferably used the variable heavy chain (VH) genes VH1-2 and VH1-69 and the variable light chain (VK) gene VK1-39 (Fig.1 b,c and Extended Data Table 3). The dominant use of these VH genes for SARS-CoV-2 S-specific antibodies is consistent with previous studies^16, 21^. In comparison, the frequency of VH1-2 and VK1-39 gene usage only ranks 23 and 6, respectively, in the B-cell repertoire of healthy individuals^22, 23^ suggesting that certain germline genes are naturally favored for binding to the S protein of SARS-CoV-2. Notably, the selected antibodies had very limited somatic hypermutation (SHM) with a median frequency of 3.2% in VH and 1.6% in VL (Fig. 1d). These findings are consistent with previous studies, which reported the isolation of anti-SARS-CoV-2 antibodies in nearly germline configuration^24–26^. The length distribution of the complementarity-determining region 3 (CDR3) in both the heavy and light chains was in line with previous observations (Fig. 1e,f), although we found an unusual bimodal distribution of CDRH3 length (15 and 20 amino acids) as opposed to the typical bell-shaped distribution in CDRL3 (9 amino acids) (Fig. 1e,f). No correlation was found between SHM frequency in VH and binding affinity (Fig. 1g), further supporting the notion that certain germline antibody genes are naturally fit for targeting the S protein.

## Potent neutralization of diverse SARS-CoV-2 variants of concern

The neutralizing activity of the selected mAbs was initially evaluated using a lentivirus-based pseudotype virus neutralization assay^27^. Eleven of the 18 mAbs showed neutralizing activity against the original SARS-CoV-2 strain pseudoviruses with an S sequence corresponding to that of an early SARS-CoV-2 isolate (WA-1). Seven of them were very potent, with half-maximal inhibitory concentration (IC50) values below 10 ng/mL (Fig. 1h), which is comparable to the potency of the best-in-class mAbs^28–30^. These 11 mAbs along with H11, which had high binding affinity for the S-2P protein by SPR, were tested for their ability to bind to the SARS-CoV-2 receptor-binding domain (RBD), the most critical target for virus neutralization, by competition ELISA using a recombinant RBD protein. Binding of all Fabs except H11 was strongly inhibited by pre-incubation with soluble RBD protein, indicating that all except H11 were specific for the RBD (Extended Data Fig. 4). To investigate if the epitopes recognized by these RBD-specific antibodies are distinct or overlapping, we performed cross-competition RBD ELISAs of these 11 antibodies using Fab fragments and complete IgG. All the antibodies showed high levels of cross-competition with the exception of two, 1G8 and 3F6, which cross-competed only between each other (Extended Data Fig. 5).

**Fig. 4.**
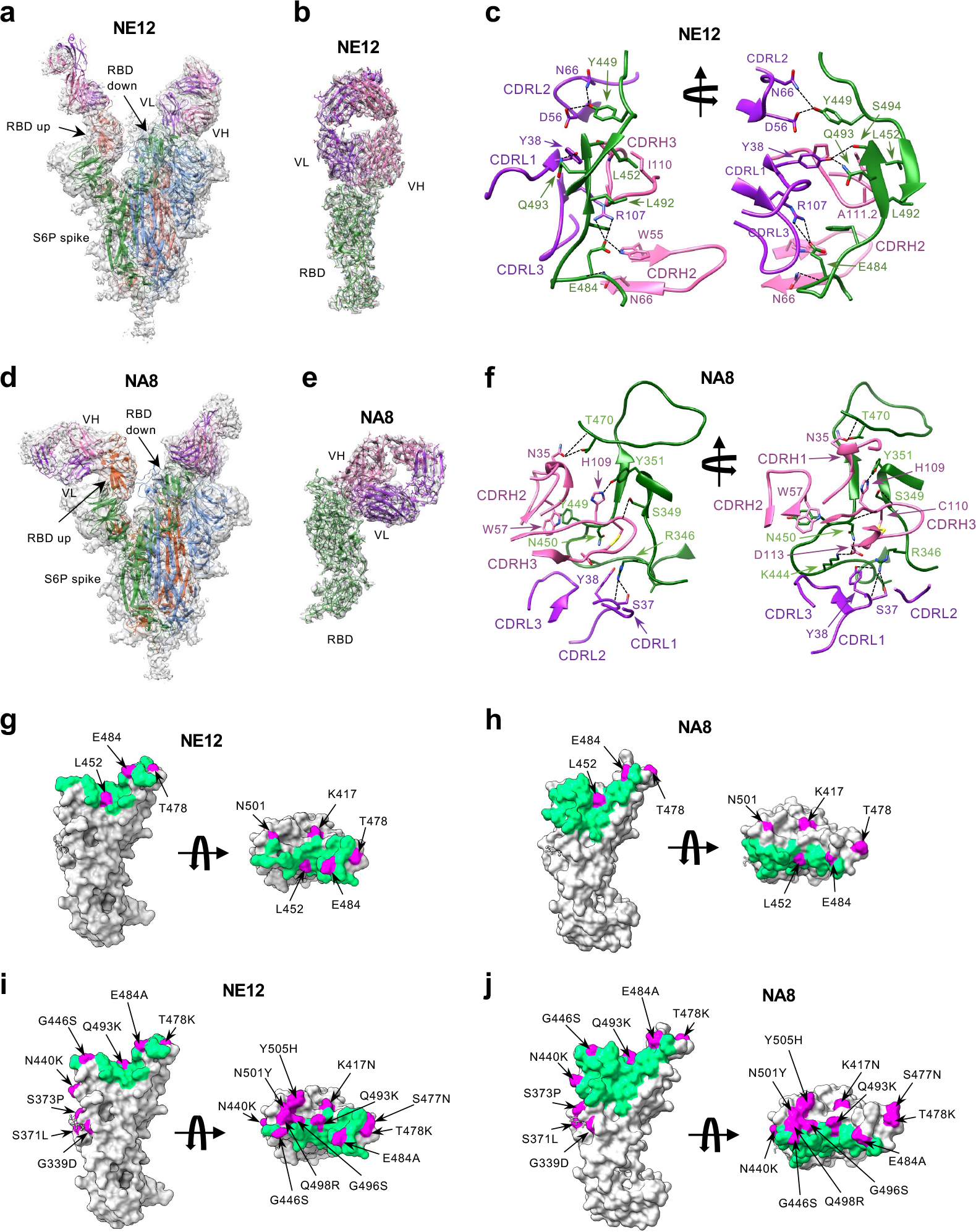
Cryo-EM structures of Fab fragments of neutralizing mAbs NE12 and NA8 in complex with a stabilized SARS-CoV-2 spike protein trimer. a, Cryo-EM map of Fab NE12 in complex with the SARS-CoV-2 S-6P spike trimer with docked spike and Fab atomic models. The heavy and light chains of the mAb are colored pink and purple, respectively. Spike protomers are colored individually. The Fab fragment binds the receptor binding domain (RBD) in both the up and down positions. b, Local refinement of a region containing one RBD with bound Fab resolved atomic details of the RBD-NE12 interactions. c, Ribbon diagram of the RBD/NE12 contact interface. Coloring is as in panel b. Only the complementarity determining regions (CDRs) and RBD fragment participating in the interaction are shown for clarity. Key residues are shown in stick representation. Dashed lines represent salt bridges and hydrogen bonds. d, Cryo-EM map of Fab NA8 in complex with the SARS-CoV-2 S-6P spike trimer with docked spike and Fab atomic models. The Fab fragment binds the receptor binding domain (RBD) in both the up and down positions. e, Local refinement of a region containing one RBD with bound Fab resolved atomic details of the RBD-NA8 interactions. f, Ribbon diagram of the RBD/NA8 contact interface. Only the CDRs and RBD fragment participating in the interaction are shown for clarity. g-j, Surface representation of the RBD with the epitopes of NE12 (g,i) and NA8 (h,j) colored in green. Residues mutated in B.1.1.7, B.1.617 and B.1.351 (g, h) or B.1.1.529/Omicron (i, j) variants of concern are colored in magenta. The Omicron mutations are labeled in panels i and j. The antibody residues are numbered according to the IMGT (http://www.imgt.org)^38^.

Next, using a pseudotype virus assay, we tested our mAbs against different VOCs of global relevance, namely, B.1.1.7/Alpha, B.1.351/Beta, B.1.617.2/Delta and B.1.1.529/Omicron, and compared their efficacy with that of three antibodies approved for clinical use, Lilly_CoV555 (Bamlanivimab)^28^, REGN10933 Casirivimab) and REGN10987 (Imdevimab)^29, 30^ (Fig. 1h). We found 7 mAbs that potently neutralized the WA-1 strain, the B.1.1.7/Alpha and B.1.617.2/Delta variants in the picomolar range. In contrast, most mAbs were ineffective against the Beta variant, B.1.351, with only four showing neutralizing activity. Remarkably one of them, NA8, neutralized B.1.351 at picomolar concentrations, with a potency similar to that of REGN10987. Impressively, the same mAb also potently neutralized the B.1.1.529/Omicron variant, containing 37 mutations within the S protein, with IC50 values in the picomolar range (Fig. 1h), whereas none of the clinically approved antibodies we tested showed neutralizing activity. Three additional mAbs (2A12, 3F6 and NE12) neutralized the Omicron variant, albeit in the nanomolar range (Fig. 1h). The neutralizing activity of our mAbs was also tested using a live virus neutralization assay against the original strain, WA-1, and the B.1.351/Beta variant (Fig. 1i). Even though this assay was less sensitive than the pseudovirus assay, the neutralization profile was similar overall, as shown by a strong correlation between the results obtained in the two assays (Fig. 1j). Importantly, our best mAbs were more potent than the clinically approved mAb Lilly_CoV555 against WA-1, with IC50 values more than 4-fold lower; furthermore, NA8 potently neutralized the B1.351 variant while the Lilly_CoV555 mAb had no activity (Fig. 1i). Based on the *in vitro* neutralization data, two mAbs, NE12 and NA8, were selected for further characterization and development. Complete neutralization curves for these two mAbs are shown in Fig.1k,l.

## Antibodies NE12 and NA8 protect hamsters from SARS-CoV-2 infection

Next, we tested the *in vivo* prophylactic efficacy of mAbs NE12 and NA8 in the golden Syrian hamster model, which closely mimics the severity of the disease in humans^31, 32^. A total of 80 male hamsters were used for these experiments; each of the study groups included 10 hamsters. The animals were inoculated with mAbs at two concentrations (12 and 4 mg/kg) via intraperitoneal (IP) injection 24 h before challenge with 4.5 log10 TCID50 of SARS-CoV-2 WA-1 or B.1.351/Beta by intranasal (IN) instillation (Fig. 2a). An isotype-matched (IgG1) irrelevant mAb, the anti-HIV-1 VRC01, was included as a control at 12 mg/kg. Based on the *in vitro* neutralization results, both NA8 and NE12 were tested against the WA-1 strain (Fig.2b,d,e), while only NA8 was tested against the B.1.351/Beta variant (Fig.2c,f,g). Animals treated with the control IgG1 lost, on average, 9% and 8% of body weight by day 5 after challenge with WA- 1 and B.1.351, respectively (Fig. 2b,c). In contrast, treatment with NE12 or NA8 significantly protected hamsters from weight loss induced by the two viral variants, although NA8 at 4 mg/kg against the B.1.351 virus showed a lesser degree of protection (Fig. 2c). The protective effect of NE12 and NA8 was confirmed by viral load measurements in nasal turbinate and lung tissues on days 3 and 5 after inoculation (n=5 per time point per group). In animals treated with NE12 or NA8 at either 12 or 4 mg/kg, the viral titers at day 3 post-infection with the WA-1 virus were significantly reduced in both tissues compared to controls; in the lungs, the virus became undetectable at day 5 post-infection (Fig.2d,e). Likewise, in animals infected with the B.1.351 variant, NA8 at both concentrations significantly reduced the viral titers in nasal turbinate and lung tissues at day 3 post-infection and completely suppressed viral replication in the lungs at day 5 (Fig.2 f,g). Overall, these data demonstrate that mAb NA8 exerted strong prophylactic protection *in vivo* from two antigenically distinct SARS-CoV-2 variants, and NE12 exerted strong prophylactic protection against SARS-CoV-2 WA-1 in a highly susceptible animal model, despite the relatively low antibody doses used.

## Antibodies NE12 and NA8 have therapeutic activity against SARS-CoV-2 in hamsters

A total of 60 golden Syrian hamsters, divided in 6 groups of 10 animals each, were utilized for a therapeutic study. Three groups of animals were infected intranasally with the original strain, WA-1, and 3 groups with the B.1.351 variant (4.5 log10 TCID50 per animal). The therapeutic efficacy of mAb NE12 or NA8 (each at 12 mg/kg) was evaluated by administering the antibodies via the IP route 24 hours after IN instillation, which is a very stringent model given the extremely rapid kinetics of replication of SARS-CoV-2 (Fig. 3a). The IgG1 isotype control mAb VRC01 (12 mg/kg) was tested in parallel as a control. Five hamsters per group were sacrificed at da y 3 post-infection for tissue viral load measurements, while the remaining 5 animals were weighed daily until day 11. No significant differences in weight loss were observed among the three groups of hamsters infected with WA-1 (Fig. 3b). In contrast, NA8 significantly reduced the weight loss caused by infection with the B.1.351/Beta variant, which remained below 5% of the initial body weight throughout the observation period in contrast with the marked loss (nearly 15%) seen in the control group (Fig. 3c). Analysis of viral titers in nasal turbinate and lung tissues showed a significant reduction in both NE12- and NA8-treated animals infected with WA-1, despite a wide data distribution in animals treated with NE12 (Fig. 3d,e). The serum concentrations of NA8 and NE12 were measured on day 3 after infection. In the NE12/WA-1 group, the antibody titers varied greatly; high viral titers in nasal turbinates and lungs strongly correlated with low to undetectable antibody levels in serum of three animals (Extended Data Fig. 6a,b). In hamsters infected with B.1.351, treatment with NA8 induced a significant reduction in viral titers in both nasal turbinate and lung tissues while, as expected, NE12 was ineffective (Fig. 3f,g). Altogether, these results demonstrate that both NE12 and NA8 exerted strong therapeutic effects against sensitive SARS-CoV-2 strains in a suitable preclinical model.

## Structural analysis of antibodies NE12 and NA8

Cryo-EM analysis was used to determine the structures of Fabs NE12 and NA8 in complex with a stabilized SARS-CoV-2 spike protein trimer (S-6P) at nominal resolutions of 3.1 Å and 2.9 Å, respectively^33^ (Fig. 4a,d, Extended Data Table 4, Extended Data Fig. 7-10). The Fab-RBD interfaces were resolved using local refinement to 3.4 Å (NE12 complex) and 3.5 Å (NA8 complex) (Fig. 4b,e, Extended Data Fig.11,12. Fab NE12 interacts with the face of the receptor-binding ridge of the RBD in both the up and down positions (Fig. 4a). The interaction buries 874 Å^2^ of the Fab surface area, split nearly equally between the heavy (465 Å^2^) and light (409 Å^2^) chains. The 17 residue-long complementarity-determining region (CDR) 3 of the heavy chain (CDRH3) and the CDR1 of the light chain (CDRL1) straddle the receptor-binding ridge, while the CDRL3 and CDRH2 interact with the tip of the RBD (Fig. 4c). The CDRH1 is disordered, while the CDRL2 interacts with the region of the receptor-binding ridge distant from the tip. The epitope of this Barnes class-2 antibody^34^ overlaps with those of mAbs Lilly_CoV555^28^ and REGN10933^29^, as well as with the ACE2 receptor footprint^35^, indicating that NE12 directly blocks the spike-receptor interaction (Extended Data Fig.13). This mechanism was confirmed by binding competition studies on ACE2-expressing cells (Extended Data Fig. 14). Residues mutated in the B.1.1.7/Alpha and B.1.617.2/Delta variants are located outside or at the periphery of the NE12 epitope, thus allowing for a potent neutralization of these variants (Fig. 4g). In contrast, the E484K mutation of B.1.351 results in clashes with CDRH2 and CDRL3 residues, compromising the neutralizing activity. Likewise, multiple replacements in the B.1.1.529/Omicron RBD receptor-binding ridge can prevent NE12-spike interactions (Fig. 4i).

The structure of Fab NA8 also shows extensive contacts with the receptor-binding ridge, but this antibody binds along the outer side of the RBD distal from the tip (Fig. 4e,f), displaying characteristics of both class-2 and class-3 antibodies^34^. Binding competition studies on ACE2- expressing cells showed evidence that NA8 reduced the interaction between the spike trimer and ACE2 by 90% (Extended Data Fig. 14). The interaction buries 981 Å^2^ of the Fab surface area, with the heavy chain responsible for most contacts (642 Å^2^). The CDRH3 forms a long loop positioned along the side of the RBD, whereas the CDRH1 and CDRH2 contact the central region of the receptor-binding ridge. The CDRL1 and CDRL2 interact with the side of the RBD, while the CDRL3 contacts the loop formed by RBD residues 444-447. Of the residues mutated in the included VOCs, only L452 is part of the NA8 epitope (Fig. 4h), which can explain the reduced neutralization of B.1.617.2/Delta (Fig. 2h). Noteworthy, all the B.1.351/Beta and B.1.1.529/Omicron mutations fall outside the NA8 epitope or are located at its periphery (Fig. 4j, Extended Data Fig.15,16), which is consistent with the high neutralization potency of NA8 against these VOCs.

## Discussion

In this study, we report the generation by combinatorial phage-display library technology of 11 potent mAbs capable of neutralizing highly diverse SARS-CoV-2 VOCs. Unlike single B-cell cloning approaches, phage-display library technology does not provide naturally paired immunoglobulin heavy and light chains and therefore opens the possibility of generating new antibodies with improved biological features. Two mAbs in particular, NE12 and NA8, represent promising candidates for clinical use. Their neutralization spectrum is different and complementary, with NE12 being extremely potent against an early SARS-CoV-2 strain, WA-1, as well as against the B.1.1.7/Alpha and B.1.617.2/Delta variants, and NA8 exhibiting an unusually broad spectrum of action with ultrapotent activity against the two most challenging current VOCs, B.1.351/Beta and B.1.529/Omicron. It should be emphasized that the B.1.351 variant is particularly difficult to neutralize and indeed very few effective mAbs are currently available. Even more striking, NA8 has picomolar neutralizing activity against the recently emerged Omicron variant, in contrast with the most widely clinically used mAbs, Lilly_CoV555^28^, REGN10933 and REGN10987^29, 30^, which have no neutralizing activity. Only one clinically approved mAb, Sotrovimab, has been reported to neutralize the Omicron variant^10, 36, 37^, albeit with markedly lower neutralization potency compared to NA8. Considering the reduced efficacy of current vaccines and the complete loss or reduced neutralizing activity by most clinically approved antibodies^10–15, 36, 37^, the identification of new antibodies capable of blocking not only the Beta but also the Omicron variant is important for future therapeutic and preventive strategies. Corroborating the potential clinical usefulness of our antibodies, the *in vitro* neutralization potency of NE12 and NA8 was confirmed in a hamster model *in vivo*, with remarkable prophylactic and therapeutic efficacy against the original WA-1 strain and the B.1.351/Beta variant at relatively low antibody dose.

Structural analysis revealed different modes of binding of NE12 and NA8 to the SARS-CoV-2 spike. Like other class-2 antibodies, which neutralize the virus by blocking the spike interaction with the ACE2 receptor, NE12 binds to the receptor-binding ridge of the RBD. In line with a similar neutralization profile, there is a substantial overlap between the NE12 epitope and that of clinically approved mAb Lilly_CoV555. In contrast, the highly conserved NA8 epitope includes a portion of the receptor-binding ridge but is shifted toward the outer side of the RBD, allowing the antibody to “sidestep” the residues that are mutated in B.1.351 and the Omicron variant. This corroborates the observation that the rare antibodies capable of neutralizing B.1.351 recognize either the inner or the outer side of the RBD^7^.

The identification of potent, broadly neutralizing antibodies against SARS-CoV-2 VOCs that resist neutralization by most mAbs, such as the B.1.351/Beta and the B.1.529/Omicron variants, is critical for creating an arsenal of therapeutic antibodies with high potency against both present and future VOCs that will continue to emerge because of the sustained worldwide spread of this virus, leading to escape from current prophylactic and therapeutic interventions.

## Data Availability

All data presented in this study are available within the article or from the corresponding authors upon request. Source data are provided with this paper. The antibody sequences reported in this paper have been deposited in the GenBank database (accession nos. OM179962-OM179997). The ensemble cryo-EM maps of the NE12/spike trimer and NA8/spike trimer complexes were deposited to the Electron Microscopy Data Bank (EMDB)

https://www.ncbi.nlm.nih.gov/genbank/

## Acknowledgements

We thank John R. Mascola, Kizzmekia S. Corbett, Nicole Doria-Rose, Lingshu Wang, Kevin Carlton and the Vaccine Production Program of the Vaccine Research Center (VRC), NIAID, NIH, for providing mAbs VRC01, Lilly_CoV555, REGN10933, REGN10987 and S306, plasmids to express the SARS-CoV-2 stabilized S-2P and S-6P spike trimers, membrane-bound spike proteins from various SARS-CoV-2 strains, the lentivirus backbone pCMV DR8.2, and human TMPRSS2; Michael Farzan for providing 293-ACE2 cells; Craig Martens for sequencing the SARS-CoV-2 stocks; and the staff of the NIAID Comparative Medicine Branch for animal study support. The authors thank Drs. Anthony S. Fauci, Steven Holland, and Jeffrey I. Cohen for critically reading the manuscript and for their helpful suggestions. This work was supported by the Intramural Research Program of the National Institute of Allergy and Infectious Diseases (NIAID), National Institutes of Health (NIH), and by the Intramural Research Program of the VRC, NIAID, NIH. This work was also supported by Federal funds from the NIAID, NIH, under Contract No. HHSN272201800013C, by Federal funds from the Frederick National Laboratory for Cancer Research (FNLCR), NIH, under Contract HHSN261200800001 (YT, TS). Cryo-EM datasets were collected at the National CryoEM Facility (NCEF) of the National Cancer Institute (NCI) with the support of the NCI National Cryo-EM Facility at the FNLCR under contract HSSN261200800001E. Chimera was developed by the Resource for Biocomputing, Visualization, and Informatics at the University of California, San Francisco, with support from NIH P41-GM103311. The Frederick Research Computing Environment (FRCE) high-performance computing cluster was used for processing cryo-EM datasets. R.G. and M.R.H. performed this work as employees of Laulima Government Solutions, LLC, while E.P performed this work as an employee of Tunnell Government Services. The content of this publication does not necessarily reflect the views or policies of the US Department of Health and Human Services (DHHS) or of the institutions and companies affiliated with the authors.

## METHODS

### Convalescent COVID-19 plasma donor selection and lymphocyte isolation

Convalescent COVID-19 plasma donors were prospectively enrolled onto an institutional review board-approved protocol (Clinical Trials Registration, NCT04360278)^19^ and provided written informed consent for the study. Among them, 12 convalescent plasma donors with high titer neutralizing antibodies against SARS-CoV2 were selected and 20-40 ml of blood was collected from each of the 12 donors. Eligibility criteria included molecular or serologic laboratory evidence of past COVID-19 infection, and complete recovery from COVID-19, with no symptoms for ≥28 days or ≥14 days with a negative molecular test after recovery. PBMCs were prepared using density centrifugation on a Ficoll-Paque gradient.

### Live virus neutralization assay used in COVID-19 convalescent patients

Neutralization of live virus by patient plasmas was tested using a fluorescence reduction neutralization assay (FRNA) with SARS-CoV-2 [2019-nCoV/USA-WA1-A12/2020 (WA-1) from the US Centers for Disease Control and Prevention, Atlanta, GA, USA] at the NIH-NIAID Integrated Research Facility at Fort Detrick, MD, USA, as previously reported^39^. In these experiments, a fixed volume of diluted virus was incubated with an equivalent volume of test plasma for 1 hour at 37 °C prior to adding the suspension to Vero E6 cells (ATCC, Manassas, VA, USA, #CRL-1586). The virus was allowed to propagate for 24 hours prior to fixing the cells. Following fixation, the cells were permeabilized and probed with a SARS-CoV-2 nucleoprotein-specific rabbit primary antibody (Sino Biological, Wayne, PA, USA, #40143- R001) followed by an Alexa594-conjugated secondary antibody (Life Technologies, San Diego, CA, USA, #A11037). The total number of infected cells in four fields per well with each field containing at least 1000 cells was quantified using an Operetta high content imaging system (Perkin Elmer, Waltham, MA, USA). Plasma was tested using two-fold serial dilutions from 1:40 to 1:1280 with four replicates per dilution. Results were calculated as the highest dilution of plasma leading to at least 50% reduction of SARS-CoV-2 titers. Each assay was controlled with internal addition of an S-specific neutralizing polyclonal antibodies. If a 1:40 dilution did not lead to at least 50% reduction of viral titer, results were reported as <1:40 even though some inhibition of virus propagation may have been present. To enable analysis of nAb kinetics over time, a value of 1:20 was used as a surrogate for results <1:40 since this value was the next serial 2-fold titer below the lowest positive result.

### Human Fab antibody library construction

Total RNA was extracted from PBMCs using RNeasy mini kit (Qiagen) and 1^st^ strand cDNA was reverse transcribed with oligo(dT) as a primer with a kit from GE Healthcare and used as a template for antibody Fab coding fragment amplification. Briefly, the γ1 heavy chain Fd (variable and first constant region) was amplified with nine human heavy chain–specific 5′ primers and a human γ1–specific 3′ primer. The κ chain was amplified by PCR with seven human κ chain–specific 5′ primers and a 3′ primer matching the end of the constant region^40^. The λ chain was amplified by PCR with six human λ chain–specific 5′ primers and a 3′ primer matching the end of the constant region^41^. PCR was performed for 30 cycles at 95 °C for1 min, 52 °C for 1 min, and 72 °C for 1 min with AmpliTaq DNA polymerase (Applied Biosystems).

Amplified κ and λ chain DNA fragments were pooled, purified, digested with SacI and XbaI. The restriction enzyme digested light chain fragments from one donor or pooled from 5 donors were then cloned into the pComb 3H vector by electroporation^42^. The recombinant plasmid DNA was introduced into *Escherichia coli* Top 10 cells (Invitrogen) by electroporation, yielding 1 × 10^7^ individual clones. The plasmid DNA containing light chain sequences was digested with XhoI and SpeI, ligated with γ1 Fd DNA cut with the same enzymes. The plasmid DNAs containing light and heavy chains were transformed into E. coli Top 10 cells by electroporation.

Four phage display Fab libraries with average size of 5 × 10^8^ individual clones for each library were generated.

### Expression and purification of recombinant SARS-CoV-2 spike protein trimers

The stabilized SARS-Cov-2 spike protein trimer (S-2P) was produced by transient transfection of 293 Freestyle cells as previously described^20^. One liter of 293 Freestyle cells at concentration of about 0.9 million cells per ml was transfected with 1 mg of S-2P plasmid, pre-mixed with 1 ml of 293fectin™ Transfection Reagent. The cells were allowed to grow for 6-7 days at 125 rpm, 37 °C, and 8 % CO2; then the supernatant was harvested by centrifugation and filtered through Millipore Sigma 0.22 um PES filter. The cleared supernatant was incubated with 5 mL of HisPur™ Ni-NTA Resin ThermoFisher for three hours, then the resin was collected and washed with PBS with 25 mM imidazole, pH 7.4. The captured SARS-CoV-2 S S-2P was eluted by PBS with 250 mM imidazole, pH 7.4. After elution, the trimeric protein was collected, concentrated and applied to a Superdex 200 16/60 gel filtration column equilibrated with PBS. Peak fractions were pooled and concentrated to 1 mg/mL. The SARS-CoV-2 S-6P trimer was expressed and purified as previously described^33^. Briefly, 1 mg of DNA encoding the S-6P spike was transfected into 293 Freestyle cells using Turbo293 transfection reagent. The cells were grown at 37°C for 6 days, after which the supernatant was harvested and applied to His-Pure affinity resin. The resin was washed with 20 mM imidazole in PBS, and the protein eluted with 20 mM HEPES, pH 7.5, 200 mM NaCl and 300 mM imidazole. The trimeric protein was further purified on a Superdex S-200 gel filtration column equilibrated in PBS and concentrated to 1 mg/ml and flash frozen in liquid nitrogen for storage at -80°C.

### Selection of specific Fab clones

For phage production, 25 ml of logarithmic bacteria culture (OD600=0.6) in 2×TY supplemented with 100 μg/mL ampicillin and 2% glucose (2×TY-Amp-Glu) were infected with M13KO7 helper phage (New England Biolabs, USA) at 7×10^9^ plaque forming unit (PFU) per ml (∼1:20 multiplicity of infection) by incubating at 37°C for 30 min without shaking, followed by 30 min at 120 rpm. Infected cells were harvested by centrifugation (5000 g for 5 min) and resuspended in 100 ml 2×TY supplemented with 100 μg/mL ampicillin and 50 μg/mL kanamycin. After overnight growth at 30°C at 250 rpm, the cells were removed by centrifugation (18000 g at 4°C for 20 min). The culture supernatant containing the phages was filtered through a 0.45-μm filter and then precipitated with 1/5 volume of 20% PEG8000 (polyethylene glycol) in a 2.5 M NaCl solution, for 1 h on ice. Phage particles were pelleted by centrifugation (9000 *g* at 4°C for 20 min) and re-dissolved in 1 mL 1×PBS.

The phage library was panned by affinity binding of Fab-expressing phages on S-2P coated on the wells of an ELISA plate. Blocking of plates and phages was conducted for 60 min using 3% skimmed milk in 1×PBS. All washing steps were performed using PBS containing 0.05% Tween20 (PBST). For each panning cycle, 1 μg/mL antigen was used to coat the polystyrene plate and after an overnight incubation, plates were washed and blocked. For the first panning cycle, approximately 1×10^11^ phages were incubated with the antigen coated plates for 2 h, followed by a total of 20 washes with PBST. The eluted phages were amplified in E. coli Top 10 cells. Bacterial culture was plated on 2×TY-Amp-Glu agar and incubated overnight at 30°C. Clones were harvested into 5 ml 2×TY-Amp-Glu and phage production for the next round of panning, which was conducted in 40 ml medium, as described above. Two additional panning cycles were repeated using the same protocol. Following three cycles of panning, the selected phages were used for infection of *E. coli*. Single colonies were randomly picked from the third cycle output and screening of specific binders was performed, using phage ELISA against S-2P, with a BSA coated-plate as a negative control.

### Sequence analysis of S-2P-positive Fab clones

We sequenced the heavy-chain variable region (VH-DH-JH genes) of the ELISA-positive SARS-CoV-2 S-2P- specific clones obtained from each library using the Applied BioSystems model 3730 automated DNA sequencer with a modified Sanger method. The closest human germline V(D)J gene segments for each unique sequence were determined using DNAPLOT with IMGT sequence database (http://www.imgt.org/IMGT). The framework and CDRs were assigned according to IMGT nomenclature. The somatic mutations were identified by comparison of V-genes (from framework 1 to 3) with the closest germline counterpart. The first 28 nucleotides corresponding to the primer sequence were not included in the analysis. The distribution of VH and JH gene usage and HCDR3 length of anti-HBc was analyzed.

### Expression and purification of Fab and IgG

The phagemid containing light chain and heavy chain was cleaved with NheI and SpeI and re-circularized after removal of the phage gene III DNA fragment from the vector to encode soluble Fab. Bacteria containing circularized DNA without phage gene III were cultured in 2×TY medium containing 2% glucose, 100 µg/mL ampicillin, and 15 µg/mL tetracycline at 30°C until the OD600 reached 0.5–1. The culture was diluted 5-fold in 2×YT medium without glucose and containing 0.2 mM isopropyl β-D-thiogalactoside (IPTG), and culture was continued at 27°C for 20 h for expression of soluble Fab. Because the Fab was tagged at the C terminus with (His)6, the expressed proteins were affinity-purified on a nickel-charged column and were further purified through a cation-exchange SP column (Cytiva).

For IgG production, Expi293F™ cells (Thermo Fisher) were transiently transfected with plasmids carrying the antibody heavy chain and light chains. Cells were grown for six days at 37°C with 8% CO2 shaking at 125 rpm according to the manufacturer’s protocol (Thermo Fisher). Cell cultures were harvested six days after transfection and centrifuged at 20,000 g for 30 min at 4°C. The supernatants were collected and filtrated through a 0.22 µm filter. The expressed IgGs were purified by affinity chromatography on an immobilized protein G column (Cytiva). The purity of the Fab and IgG was evaluated by sodium dodecyl sulfate-polyacrylamide gel electrophoresis (SDS-PAGE), and the protein concentrations were determined by optical density (OD) measurements at 280 nm, with an *A*280 of 1.5 corresponding to 1.0 mg/mL.

### ELISA assays

For conventional antigen-binding ELISA, 96-well plates were coated with 100 µl/well containing 1 µg/ml protein in 1×PBS (pH 7.4) and incubated at room temperature overnight. Serial dilutions of soluble Fab, IgG, or phage were added to the wells, and plates were incubated for 2 h at room temperature. The plates were washed, and the secondary conjugated antibody (anti-His-HRP, anti-human Fab-HRP, or anti-M13-HRP) was added and incubated for 1 h at room temperature. The plates were washed, and the color was developed by adding tetramethylbenzidine reagent (KPL; Gaithersburg, MD), and the development was stopped with H2SO4 after 10 min. The plates were read at OD450 in an ELISA plate reader. The data were plotted, and the dose-response curves were generated with Prism software (Graphpad Software, Inc., San Diego, CA).

For RBD-competition ELISA, 96-well plates were coated with 100 µl/well containing 1 µg/mL of recombinant S-2P trimer in 1×PBS (pH 7.4), and the plates were incubated at room temperature overnight. Each Fab was 3-fold serially diluted from a starting concentration of 3 µg/mL in 3% milk/PBS or in 3% milk/PBS containing recombinant RBD protein (Sino Biological #40592-V08B) at fixed concentration (5 µg/mL). Following incubation for 2 h at room temperature, the plate was washed, and a secondary anti-human Fab-HRP antibody (Jackson ImmunoResearch) was added for 1 h at room temperature. The plate was then washed, and the reaction was developed by adding tetramethylbenzidine substrate (KPL; Gaithersburg, MD). The reaction was stopped after 10 min by addition of sulfuric acid. The plates were read at OD450 in an ELISA plate reader. The data were plotted, and dose-response curves with and without inhibition with RBD were generated using Prism software (Graphpad Software, Inc., San Diego, CA).

For antibody cross-competition ELISA, 96-well ELISA plates were coated with 100µl/well containing 1 µg/mL of recombinant S-2P trimer in 1×PBS (pH 7.4) and the plates were incubated at room temperature overnight. To test cross-competition among the mAbs, the IgG version of each mAb was incubated at 0.1 µg/ml with each of 11 Fabs at 1 µg/ml. An in house-produced anti-poliovirus IgG antibody was used as a negative control^43^. Following incubation for 2 h at room temperature, the plates were washed, and a secondary anti-human IgG Fc-HRP antibody (Jackson ImmunoResearch) was added and incubated for 1 h at room temperature. The plates were then washed, and the reaction was developed by adding tetramethylbenzidine substrate (KPL; Gaithersburg, MD). The reaction was stopped after 10 min by addition of sulfuric acid. The plates were read at OD450 in an ELISA plate reader. The average value from three wells was used to calculate the percent competition using the following formula: [1- (average OD from wells containing test IgG with Fab – average OD from control wells)/(average OD from wells containing test IgG without Fab – average OD from control wells)] x100%.

### Surface plasmon resonance (SPR)

The SPR experiments were performed on a Biacore™ T200 (Cytiva) at 25 °C in 10 mM HEPES pH 7.2, 150 mM NaCl, 3 mM EDTA, 0.05% Tween-20. The recombinant SARS-CoV-2 spike 2p (S-2P) protein was immobilized on a Series S-CM5 sensor chip (Cytiva) by amine coupling (NHS/EDC) to flow cells. A mock coupled surface was used for background subtraction.

Binding and kinetics studies were performed multiple times for each Fab. Increasing amounts of each Fab were injected over the surface at a flow rate of 30 μl/min with an association time of 120 sec and dissociation time of 180 sec, followed by a regeneration with 100mM CAPS, pH 11.5 for 90 sec. Binding data were analyzed using Biacore™ T200 Evaluation Software 3.1 (Biaeval) with fits to the Langmuir binding equation for a 1:1 interaction model. In addition, data were analyzed by surface site affinity distribution analysis by EVILFIT^44^. Values obtained with the two methods were consistent.

### Pseudotype neutralization assay

The SARS-Cov-2 pseudovirus neutralization assays were performed as previously reported^45^. Briefly, single-round luciferase-expressing pseudoviruses were generated by co-transfection of plasmids encoding the full-length SARS-CoV-2 S protein (Wuhan-1, GenBank accession number, MN908947.3; B.1.351/Beta; B.1.1.7/Alpha; B.1.617.2./Delta; B.1.1.529/Omicron; luciferase (pHR’ CMV Luc), a lentivirus backbone (pCMV ΔR8.2), and human transmembrane protease serine 2 (TMPRSS2) at a ratio of 1:20:20:0.3 into HEK293T/17 cells (ATCC) using the transfection reagent LiFect293™. The pseudoviruses were harvested after 72 hours. The supernatants were collected by centrifugation at 1500 rpm for 10 minutes, then filtered through a 0.45 µM filter, aliquoted and titrated before the neutralization assay. To test antibody-mediated neutralization, an 8-point, 5-fold dilution series was prepared for each antibody in culture medium (DMEM medium supplemented with 10% fetal bovine serum, 1% penicillin-streptomycin and 3 µg/mL puromycin). Each antibody dilution (50 µL) was mixed with 50 µL of diluted pseudovirus in 96-well plates and incubated for 30 min at 37°C. The mixture was then incubated with 10^4^ ACE-2-expressing 293T cells (293T-hACE2.MF) in a final volume of 200 µL. 72 hours later, the supernatant was carefully removing, the cells were lysed with Bright- Glo™ Luciferase Assay substrate (Promega), and the luciferase activity (relative light units, RLU) was measured. Percent neutralization was normalized considering uninfected cells as 100% neutralization and cells infected in the absence of antibodies as 0% neutralization. IC50 titers were determined using a log (agonist) vs. normalized response (variable slope) nonlinear function in Prism v8 (GraphPad).

### Live Virus Neutralization Assay of hamster sera

SARS-CoV-2 neutralization using live virus was determined in the BSL3 laboratory using SARS-CoV-2 USA-WA1/2020 (WA-1; GenBank MN985325; GISAID: EPI_ISL_404895; obtained from Dr. Natalie Thornburg, Centers for Disease Control and Prevention (CDC)) and USA/MD-HP01542/2021 (lineage B.1.351/Beta variant; GISAID: EPI_ISL_890360; obtained from Dr. Andrew Pekosz, Johns Hopkins University). WA-1 was passaged twice on Vero E6 cells. The USA/MD-HP01542/2021 was passaged on TMPRSS2-expressing Vero E6 cells. The SARS-CoV-2 stocks were titrated in Vero E6 cells by determination of the 50% tissue culture infectious dose (TCID50) as previously described^46^.

The mAbs were serially diluted in Opti-MEM and mixed with an equal volume of SARS-CoV-2 (100 TCID50) and then incubated at 37°C for 1 h. Mixtures were added to quadruplicate wells of Vero E6 cells in 96-well plates and incubated for four days. The 50% neutralizing dose (ND50) was defined as the highest dilution of serum that completely prevented cytopathic effect in 50% of the wells and was expressed as a log10 reciprocal value^47^. The dilution of serum that completely prevented cytopathic effect in 50% of the wells (ND50) was calculated by the Reed and Muench formula^48^.

### Prophylactic and therapeutic studies in the hamster model

The preclinical hamster studies were approved by the NIAID Animal Care and Use Committee. All the animal experiments were carried out following the Guide for the Care and Use of Laboratory Animals by the NIH. Golden Syrian hamsters (*Mesocricetus auratus*, Envigo Laboratories, Frederick, MD) were used in experiments conducted in BSL2 and BSL3 facilities approved by the USDA and CDC. In the prophylactic study, 80 male Syrian hamsters, aged 8-9 weeks, were randomly divided into groups of 10 animals each, bled for baseline serology and inoculated intraperitoneally with 0.5 mL of NE12, NA8, or IgG1 control at 12 mg/kg or 4 mg/kg, diluted to the concentration of 2.4 mg/mL or 0.8 mg/mL in PBS. Twenty-four hours later, the animals were challenged intranasally with 4.5 log10 TCID50 of SARS-CoV-2 WA-1 or B.1.351 in 100 µL volumes of L-15 medium (ThermoFisher) per animal. Intranasal instillations were performed under light isoflurane anesthesia. Animals were euthanized by CO2 inhalation prior to necropsy. Body weights and clinical symptoms were monitored daily from day -3 before challenge through day 5 after challenge. On days 3 and 5 post-challenge, 5 animals per group were necropsied; nasal turbinate and lung tissues were collected. Tissues were weighed, mixed with L-15 medium (10 mL per gram of tissue), homogenized, and clarified by centrifugation. Aliquots were snap-frozen and stored at -80°C. The presence of the challenge virus in clarified tissue homogenates was evaluated by limiting-dilution titrations on Vero E6 cells. The titration of SARS-CoV-2 was performed by determination of the TCID50 in Vero E6 cells and expressed as TCID50 per gram of tissue, as previously described^46^.

In the therapeutic study, 60 male Syrian hamsters, aged 9-10 weeks, were randomly divided into three groups of 10 animals each, bled for serology and inoculated intranasally with 4.5 log10 TCID50 of SARS-CoV-2 WA-1 or B.1.351 in 100 µL volumes per animal. Twenty-four hours later, the animals were inoculated intraperitoneally with 0.5 mL of NE12, NA8, or IgG1 control at 12 mg/kg, diluted to the concentration of 2.4 mg/mL in PBS. Body weights and clinical symptoms were monitored daily from day -4 before virus inoculation through day 11 after inoculation. On day 3 post-inoculation, five animals per group were necropsied; nasal turbinates and lungs were collected and processed as above. Virus titers were determined by limiting-dilution titrations on Vero E6 cells as described above.

### Statistical analysis

Data sets were assessed for significance using one-way ANOVA with Tukey’s multiple comparison test, or, if indicated, by non-parametric Kruskal-Wallis test with a Benjamini, Krieger and Yekutieli false-discovery test. Statistical comparisons of weight changes between treatment groups were performed using a repeated-measures mixed-effects model with Tukey’s multiple-comparison test. Data were only considered significant at p ≤ 0.05. Analyses were performed using Prism 9 (GraphPad Software).

### Cryo-EM specimen preparation and data collection

SARS-CoV-2 S-6P spike at a concentration of 0.5 mg/ml in PBS was mixed with Fab NA8 or Fab NE12 using a Fab-to-RBD molar ratio of 1:1, and the complex was used for specimen preparation after a brief (5-10 min) incubation at 4°C. To prepare cryo-EM specimens, Quantifoil R 2/2 gold grids were glow-discharged using a PELCO easiGlow device (air pressure: 0.39 mBar, current: 20 mA, duration: 30 s) immediately before vitrification using an FEI Vitrobot Mark IV plunger. The Vitrobot chamber was kept at 4°C and 95% humidity, and the drop volume was 2.7 µl. Datasets were collected at the National CryoEM Facility (NCEF), National Cancer Institute, using a Thermo Scientific Titan Krios G3 electron microscope equipped with a K3 direct electron detector and Gatan Quantum GIF energy filter (slit width: 20 eV) (Extended Data Table 4).

### Single particle analysis of cryo-EM data

Single particle analysis was performed using the Frederick Research Computing Environment (FRCE) computing cluster. MotionCorr2 was used for patch-based movie frame alignment^49^. Contrast transfer function parameters were estimated with ctffind 4.1^50^. The following steps were performed using Relion 3.1.0^51^, unless otherwise stated. Particle picking was performed using the template-free Laplacian-Gaussian filter-based approach. Particles were first extracted with 4x binning and subjected to 2D and 3D classification, with the low-pass filtered cryo-EM map of individual SARS-CoV-2 S-2P spike at pH 7.4 serving as the initial 3D model. Particles contributing to high-quality 3D classes were then re-extracted with 2x binning, and the above procedure was repeated. Finally, the best particles were re-extracted without binning, and the 3D map was iteratively improved with rounds of 3D auto-refinement, CTF refinement, and Bayesian polishing. For local refinement, a soft mask encompassing one RBD in the down position with the bound Fab and the neighboring NTD was created after segmenting the refined ensemble map in UCSF Chimera^52^ and used for auto-refinement with limited angular search (-sigma_ang = 5). Resolution was calculated using the gold-standard approach^53^ at the FSC curve threshold of 0.143. Local resolution was determined with ResMap 1.1.4^54^.

### Atomic model generation

An RBD from the cryo-EM structure of the S-2P spike at pH 4.0 (PDB ID 6xlu) was docked into the locally refined cryo-EM map, along with an NTD from the same structure and a homology model of the corresponding Fab prepared using the SWISS-MODEL server^55^. The atomic model was refined by alternating rounds of model building in Coot^56^ and real-space refinement in Phenix^57^. Structure validation was performed with Molprobity^58^. Map-model correlations were evaluated with phenix.mtriage^59^. Molecular graphics and analyses were performed with UCSF Chimera^60^.

## Author Contributions

Z.C. and P.F conceived the research and designed the study. Z.C., P.Z., Y.M., Y.T., D.H.M., P.D.K., H.J.A., U.J.B., P.L., and P.F designed experiments. K.W., V.D.G., H.J.A contributed to donor recruitment and blood sample collection. Z.C. generated Fab libraries and cloned monoclonal antibodies. H.N. and A.P. contributed to plasmid DNA preparation and tissue culture for IgG production. P.Z. and P.L. produced pseudoviruses and performed pseudovirus neutralization assays. E.P., R.G., M.R.H., Y.M., and C.S performed live virus neutralization assays. L.F.B. and D.H.M performed binding assays and data analysis. Y.M. and C.S, performed animal studies, and Y.M. and U.J.B. analyzed results. N.L.G. and R.F. J. generated and characterized the SARS-CoV-2 stocks for the animal studies. A.O. expressed, purified, and provided S-6P spike for cryo-EM structural analysis. T.S. carried out specimen preparation for cryo-EM experiments. Y.T. performed cryo-EM experiments and with P.D.K. performed data analysis and interpretation of the cryo-EM structures. P.F. with Z.C., Y.T., U.J.B. and P.L. wrote the manuscript with all authors providing revisions and comments. P.F. provided supervision for the entire project.

## Data availability

All data presented in this study are available within the article or from the corresponding authors upon request. Source data are provided with this paper. The antibody sequences reported in this paper have been deposited in the GenBank database (accession nos. OM179962-OM179997).

The ensemble cryo-EM maps of the NE12/spike trimer and NA8/spike trimer complexes were deposited to the Electron Microscopy Data Bank (EMDB) with accession numbers EMD-XXXX and EMD-XXXX, respectively. The locally refined maps of the NE12/spike trimer and NA8/spike trimer complexes were deposited to EMDB with accession numbers EMD- XXXX and EMD-XXXX, respectively, and the corresponding coordinates were deposited to the Protein Data Bank with accession numbers YYYY and YYY.

---

**Extended Data Figure 1.**
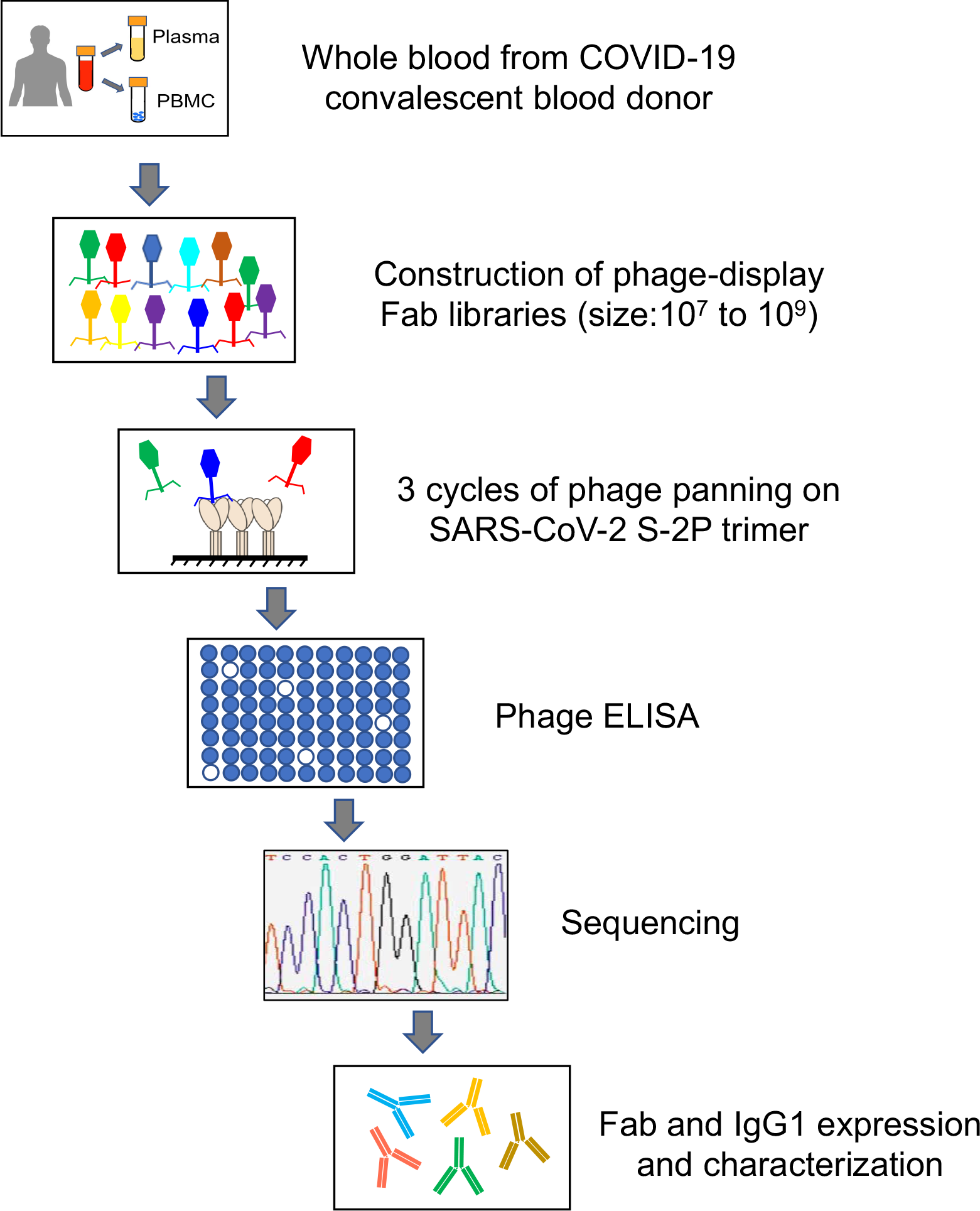
Design of the study. Schematic representation showing the different phases of phage display technology leading to the development of SARS-CoV-2 neutralizing antibodies (mAbs) starting with whole blood from 12 COVID-19 convalescent donors.

**Extended Data Figure 2.**
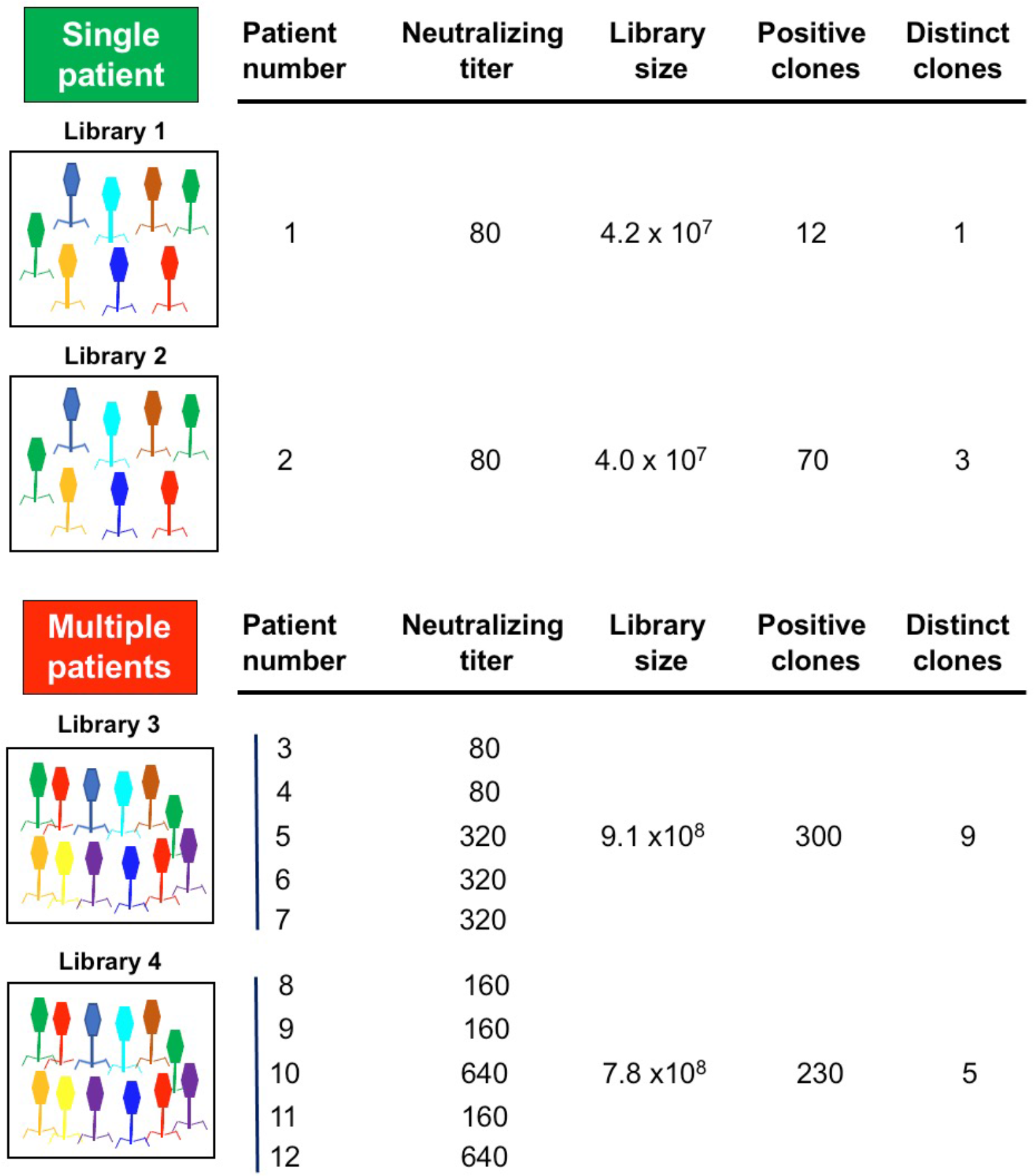
Characteristics of 4 phage-display Fab libraries. The libraries were generated from peripheral blood mononuclear cells (PBMC) obtained from convalescent COVID-19 donors, derived from either a single donor or five donors combined.

**Extended Data Figure 3.**
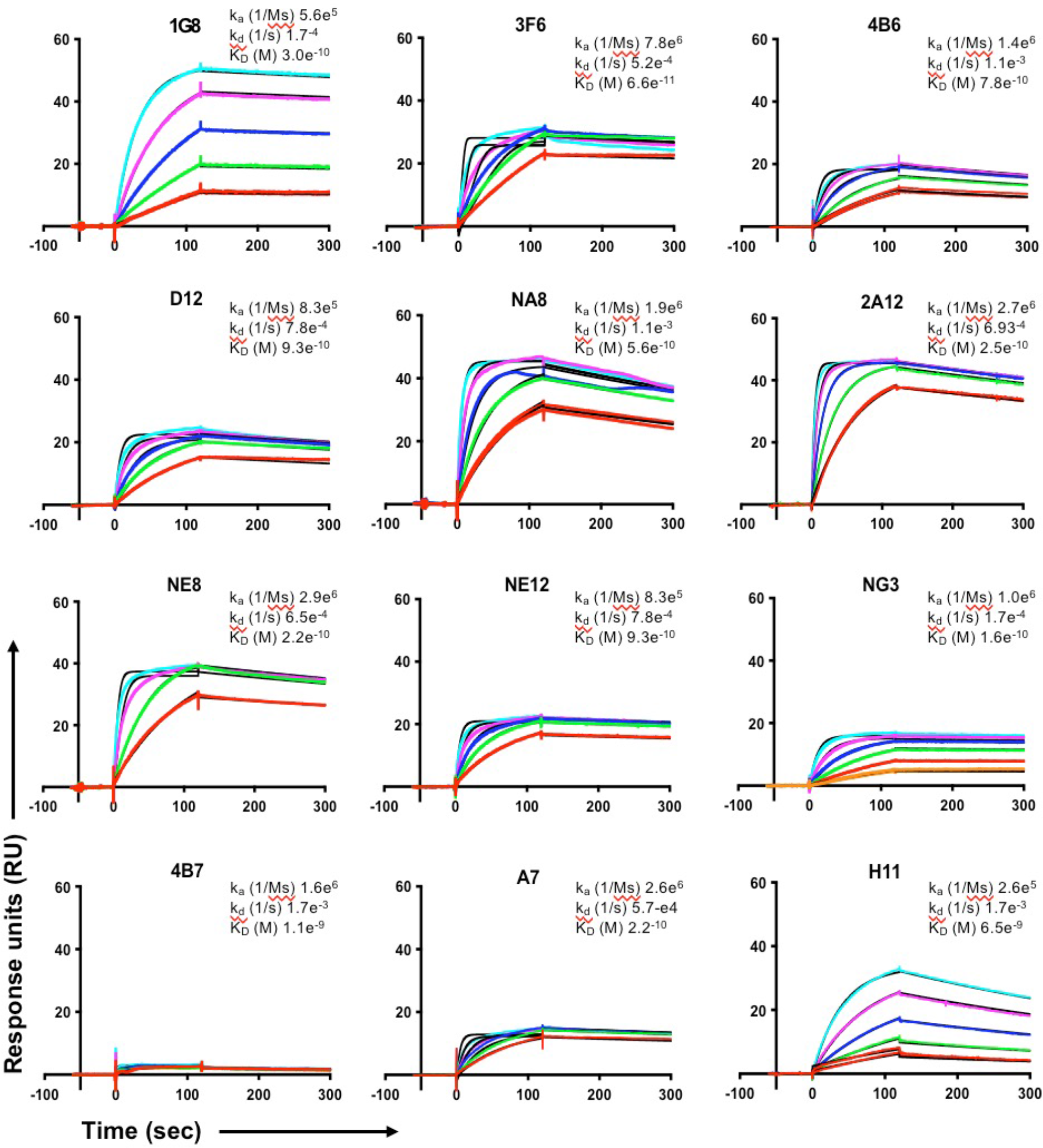
Binding profiles of 12 Fabs to a stabilized trimeric SARS- CoV-2 spike protein (S-2P). All the Fabs showed tight binding to the protein. The S-2P protein was coupled to the chip at a surface density 200-450 Rus. Fabs were flowed over the surface at graded concentrations ranging from 5.625 nM to 90 nM (color coded). Kinetic values were obtained using Biaeval 3.1; fits are indicated by the black lines.

**Extended Data Figure 4.**
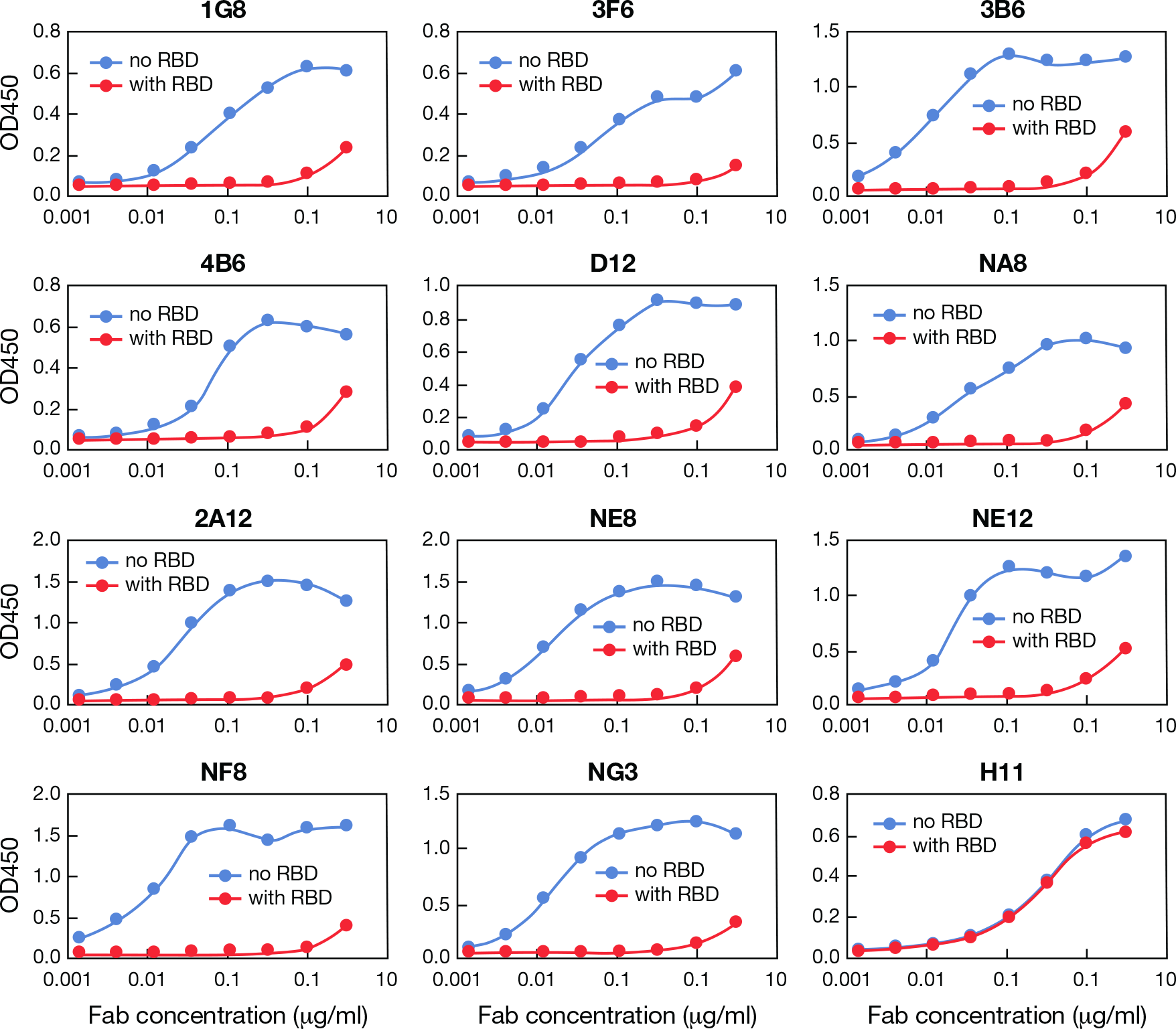
Binding of 12 Fabs derived from convalescent COVID-19 donors to the receptor-binding domain (RBD) of the SARS-CoV-2 spike protein. All of the Fabs shown, except one, are specific for the RBD, as demonstrated using the recombinant RBD protein as a competitor in ELISA. The orange lines show the loss of signal when recombinant RBD was pre-incubated with the Fabs. Only one Fab, H11, was not inhibited, suggesting that its binding site is outside the RBD.

**Extended Data Figure 5.**
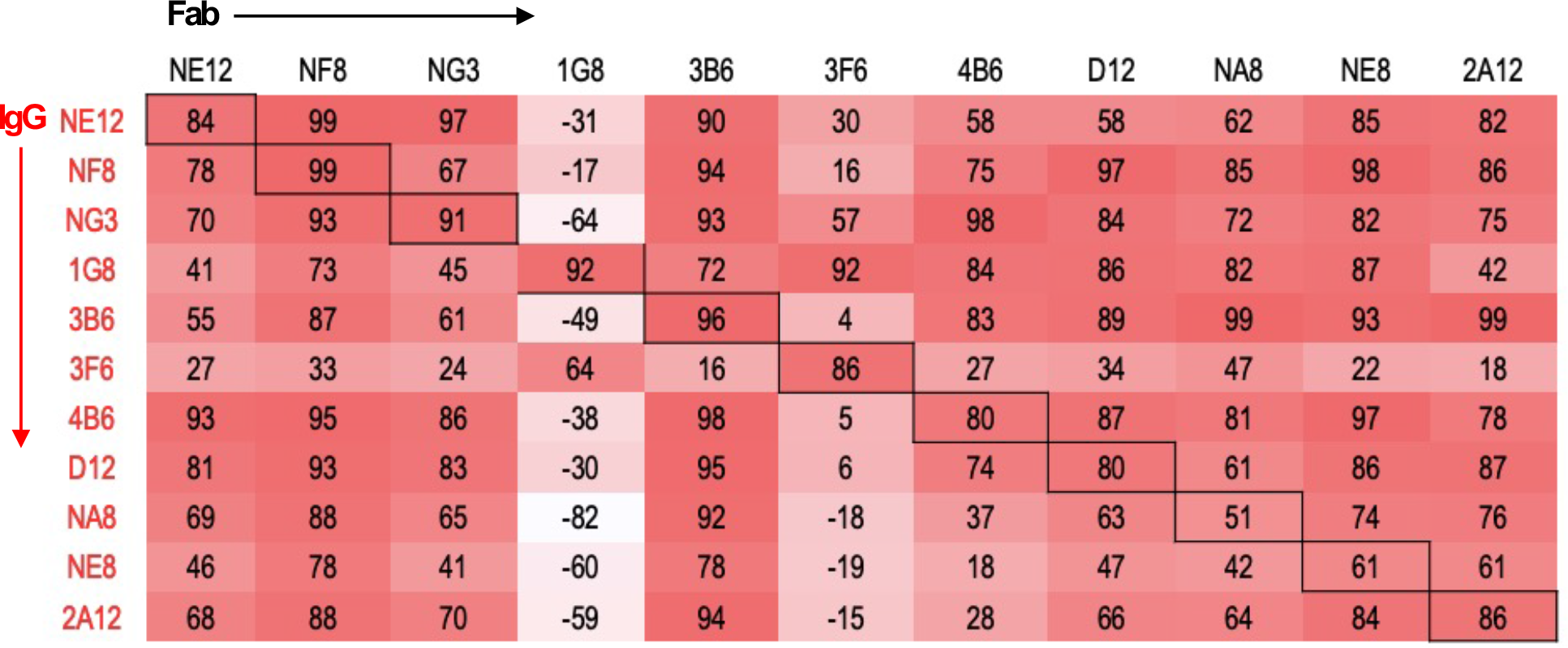
Cross-competition of 11 monoclonal antibodies for binding to the SARS-CoV-2 spike protein (S-2P) trimer. To investigate if the epitopes recognized by the 11 monoclonal antibodies are distinct or overlapping, we performed cross-competition experiments between Fabs and complete IgG in ELISA. The percent competition was calculated using the following formula: [1- (average OD from wells containing test IgG with Fab – average OD from control wells)/(average OD from wells containing test IgG without Fab – average OD from control wells)] x100%.

**Extended Data Figure 6.**
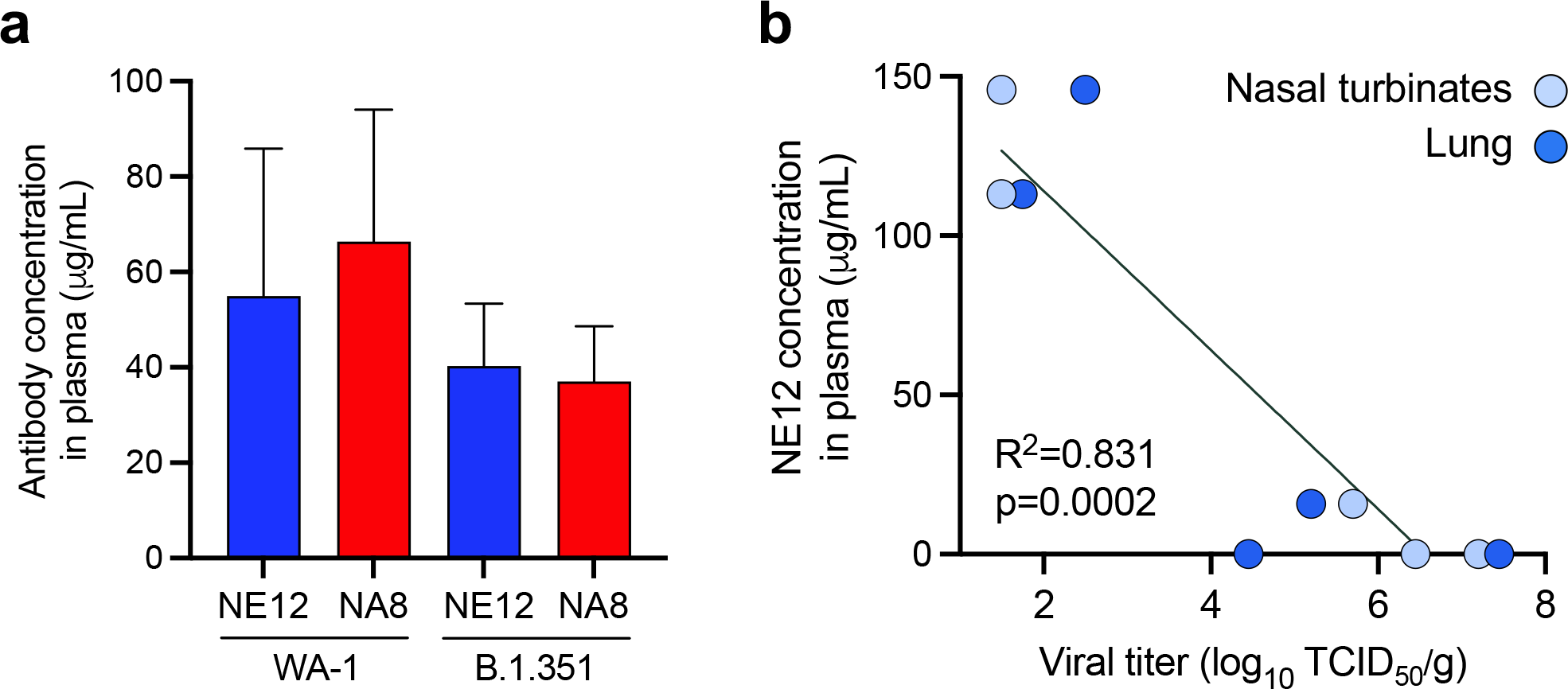
Plasma concentrations of anti-SARS-CoV-2 spike human antibodies in hamsters treated with mAbs NE12 and NA8. **a,** Mean plasma levels of NE12 and NA8 in hamsters treated with the mAbs 24 hours after virus challenge with the original strain, WA-1, or the B.1.351 variant. The mAb concentrations were determined using a specific ELISA with the recombinant S- protein trimer (S-2P) immobilized on the plastic surface. The data represent mean values from 5 animals (± SEM). **b,** Linear correlation between plasma levels of mAb NE12 and viral titers measured at day 3 in the nasal turbinate (light blue) and lung (dark blue) tissues, as determined by Pearson’s correlation.

**Extended Data Figure 7.**
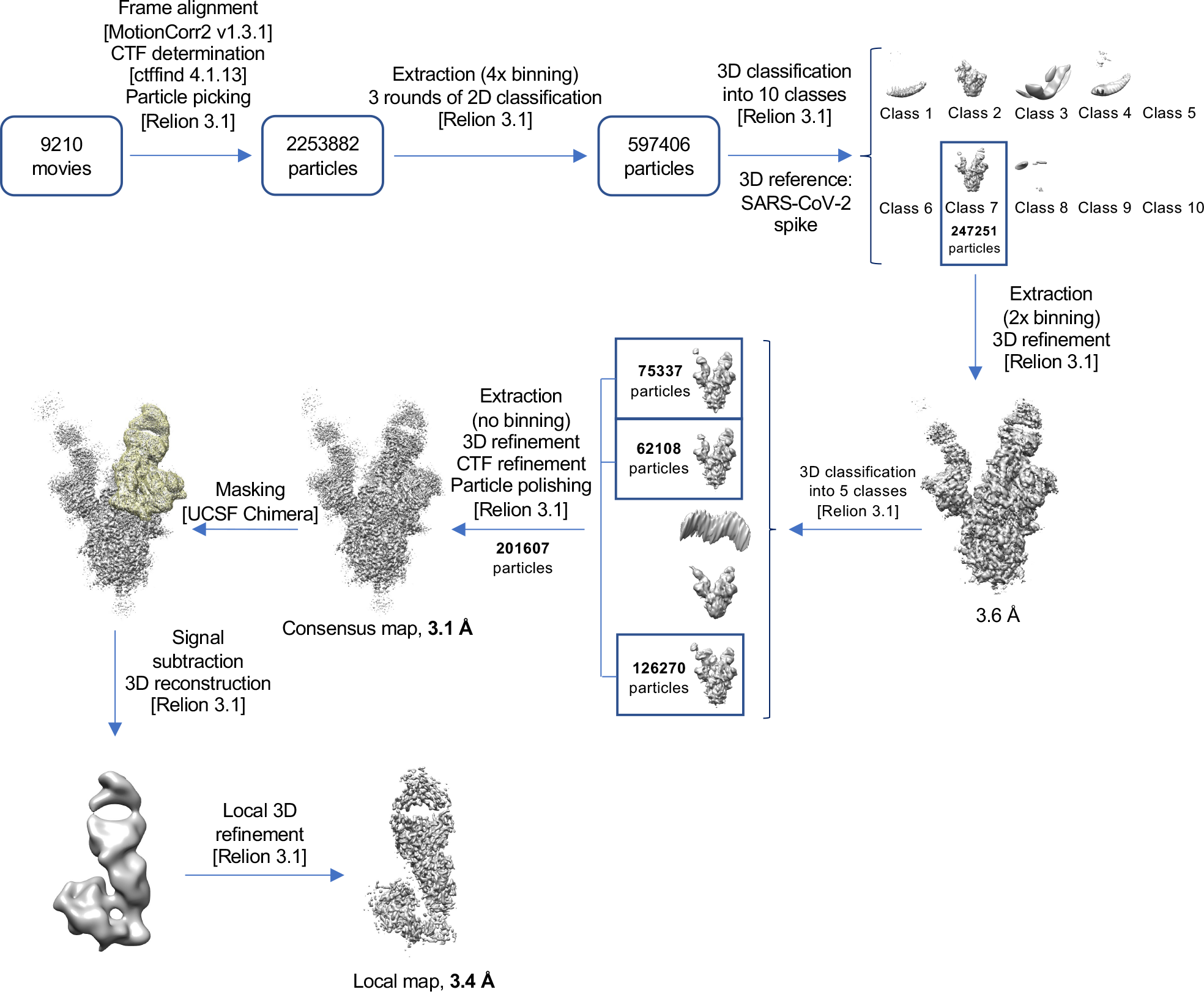
Cryo-EM data processing workflow leading to the structure of SARS-CoV-2 Spike in complex with Fab NE12. Software packages are indicated in square brackets.

**Extended Data Figure 8.**
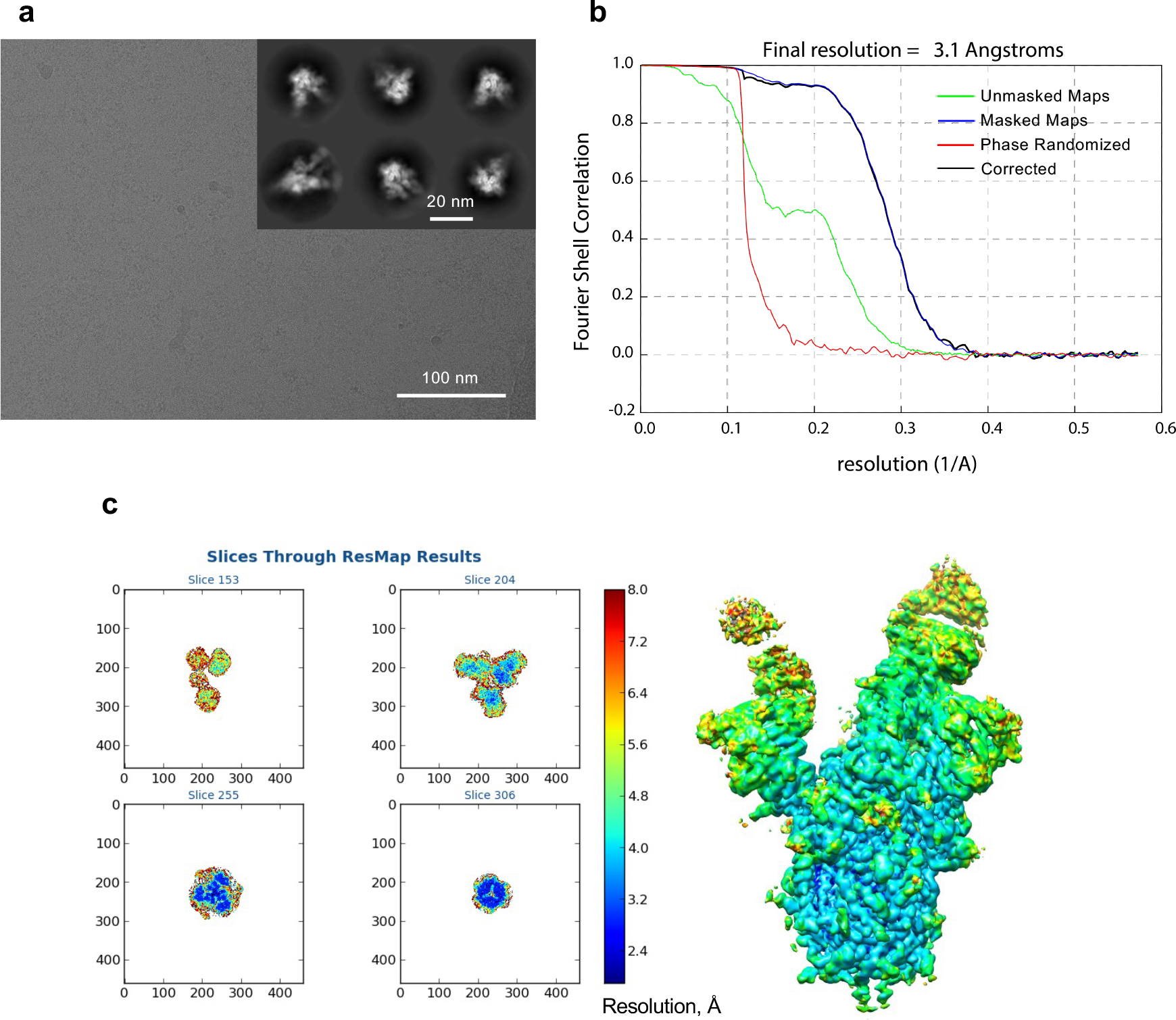
Validation of the consensus cryo-EM map of SARS- CoV-2 spike in complex with Fab NE12. **a,** Representative micrograph and 2D classes (insert). **b,** Fourier shell correlation (FSC) plots generated by Relion. **c,** Local resolution determined with ResMap.

**Extended Data Figure 9.**
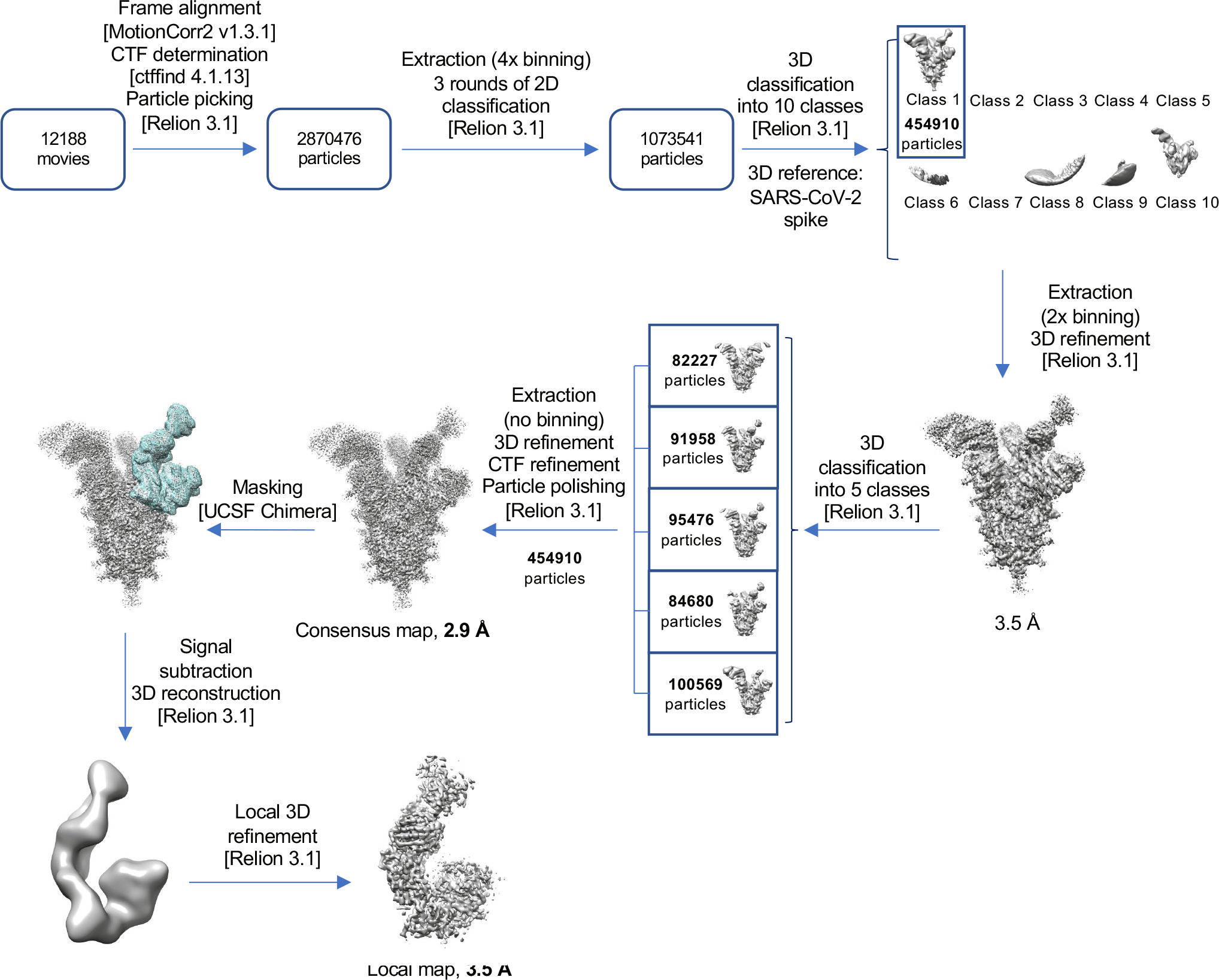
Cryo-EM data processing workflow leading to the structure of SARS-CoV-2 Spike in complex with Fab NA8. Software packages are indicated in square brackets.

**Extended Data Figure 10.**
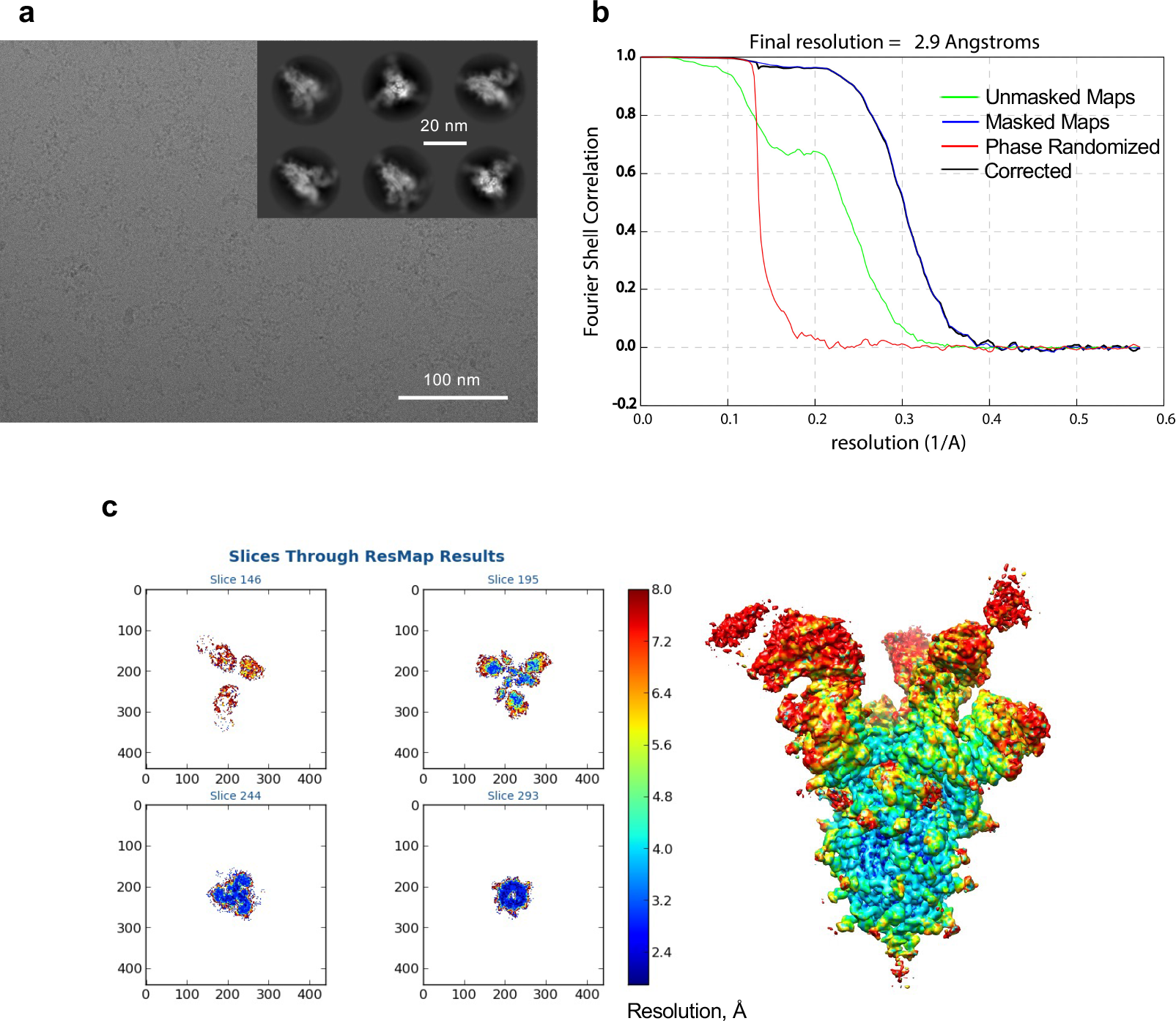
Validation of the consensus cryo-EM map of SARS- CoV-2 spike in complex with Fab NA8. **a,** Representative micrograph and 2D classes (insert). **b,** Fourier shell correlation (FSC) plots generated by Relion. **c,** Local resolution determined with ResMap.

**Extended Data Figure 11.**
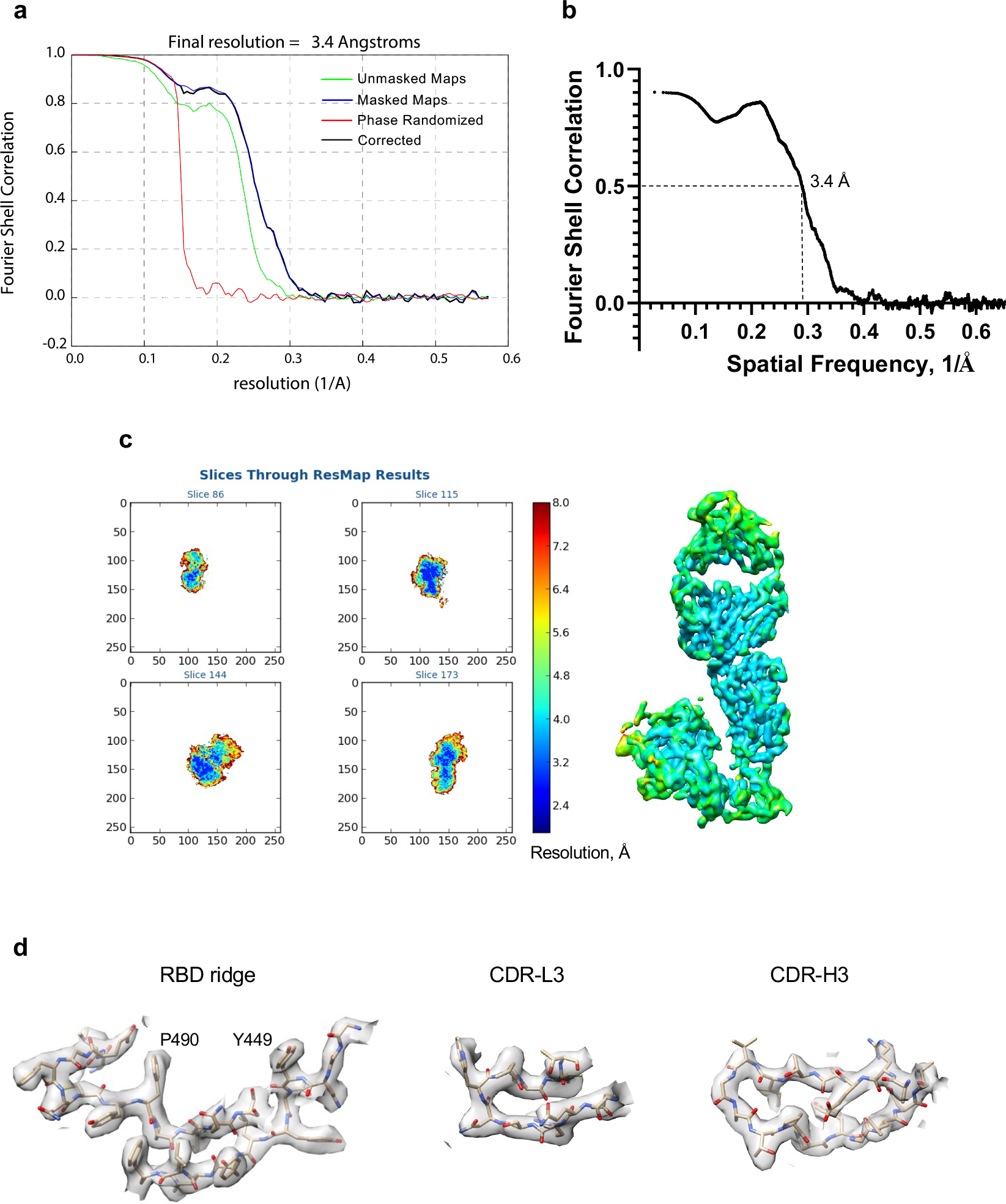
Validation of the local cryo-EM structure of SARS-CoV-2 receptor binding domain (RBD) in complex with Fab NE12. **a,** Fourier shell correlation (FSC) plots generated by Relion. **b**, FSC curve between the map and atomic model. **c,** Local resolution determined with ResMap. **d,** Examples of cryo-EM density.

**Extended Data Figure 12.**
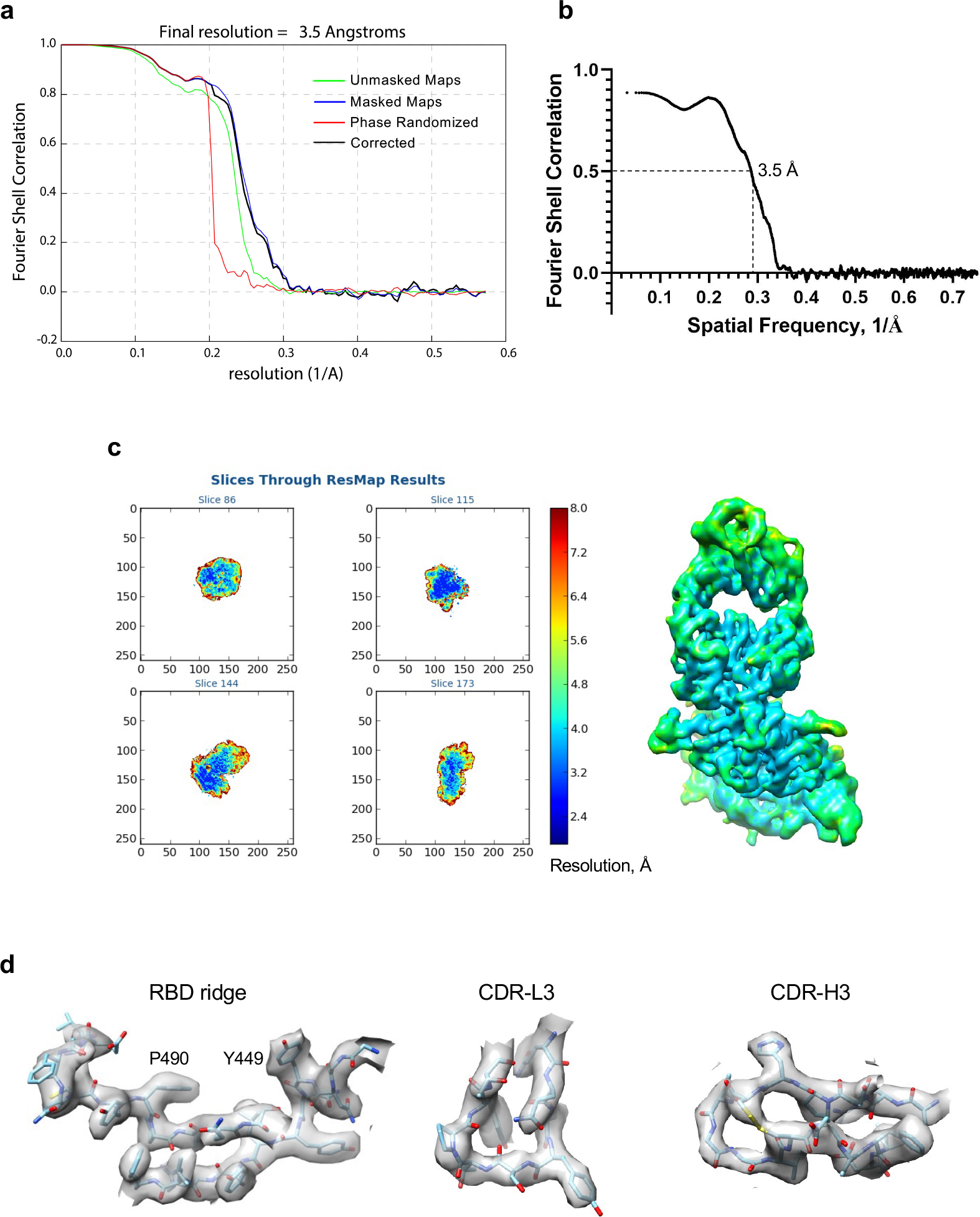
Validation of the local cryo-EM structure of SARS-CoV- 2 receptor binding domain in complex with Fab NA8. **a,** Fourier shell correlation (FSC) plots generated by Relion. **b,** FSC curve between the map and atomic model. **c,** Local resolution determined with ResMap. **d,** Examples of cryo-EM density.

**Extended Data Figure 13.**
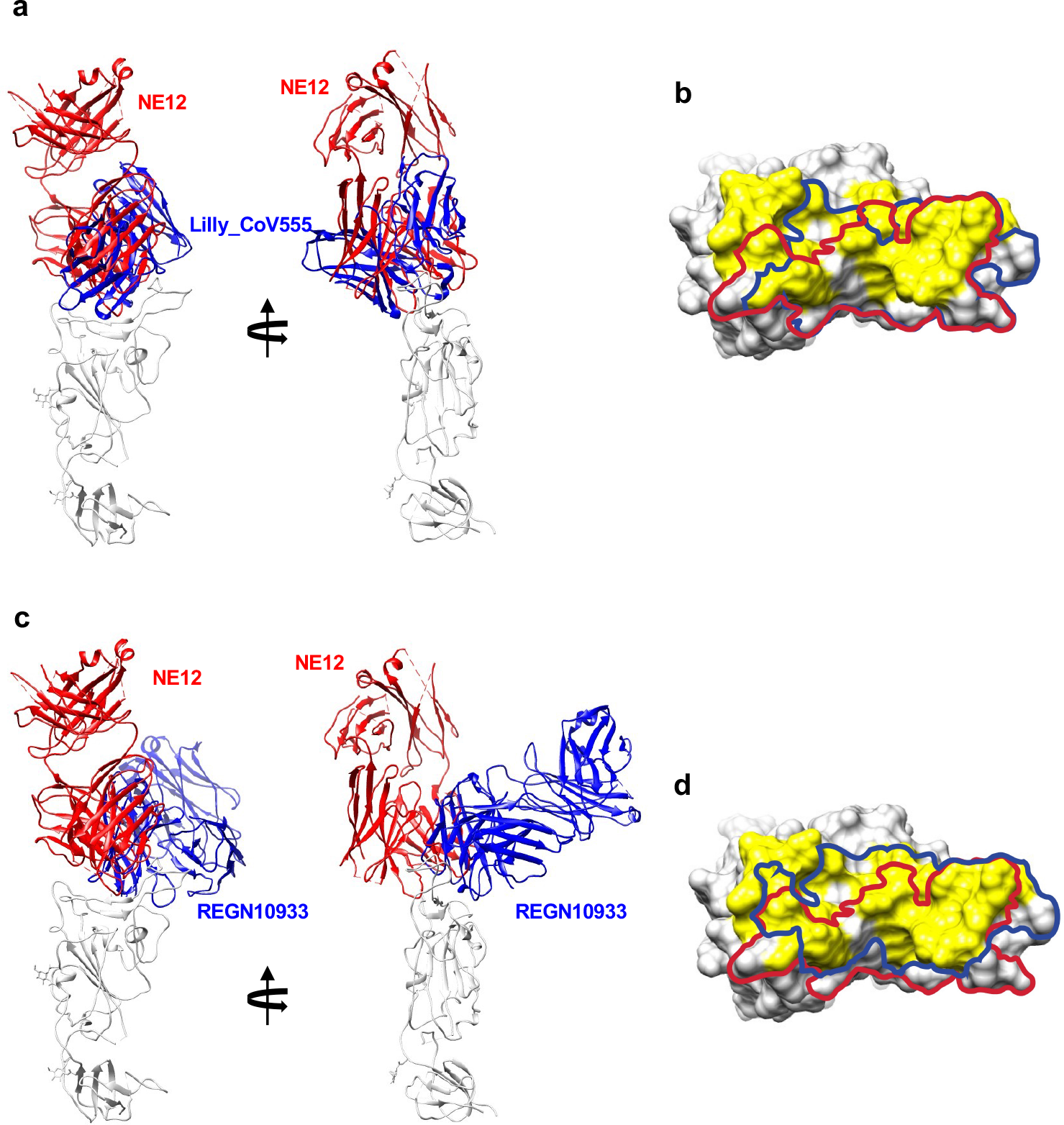
**Comparison of binding modes and epitopes of RBD- directed antibodies NE12, Lilly_CoV555 and REGN10933. a**, Cartoon representations of aligned RBD-Fab complexes for NE12 and Lilly_CoV555 (from PDB 7l3n). **b**, The RBD is shown in surface representation with the epitopes of NE12 and Lilly_CoV555 outlined with red and blue curves, respectively. Surface areas corresponding to the RBD residues forming the ACE-2 receptor interface are colored yellow. **c**, Cartoon representations of aligned RBD-Fab complexes for NE12 and REGN10933 (from PDB 6xdg). **d**, The RBD is shown in surface representation with the epitopes of NE12 and REGN10933 outlined with red and blue curves, respectively.

**Extended Data Figure 14.**
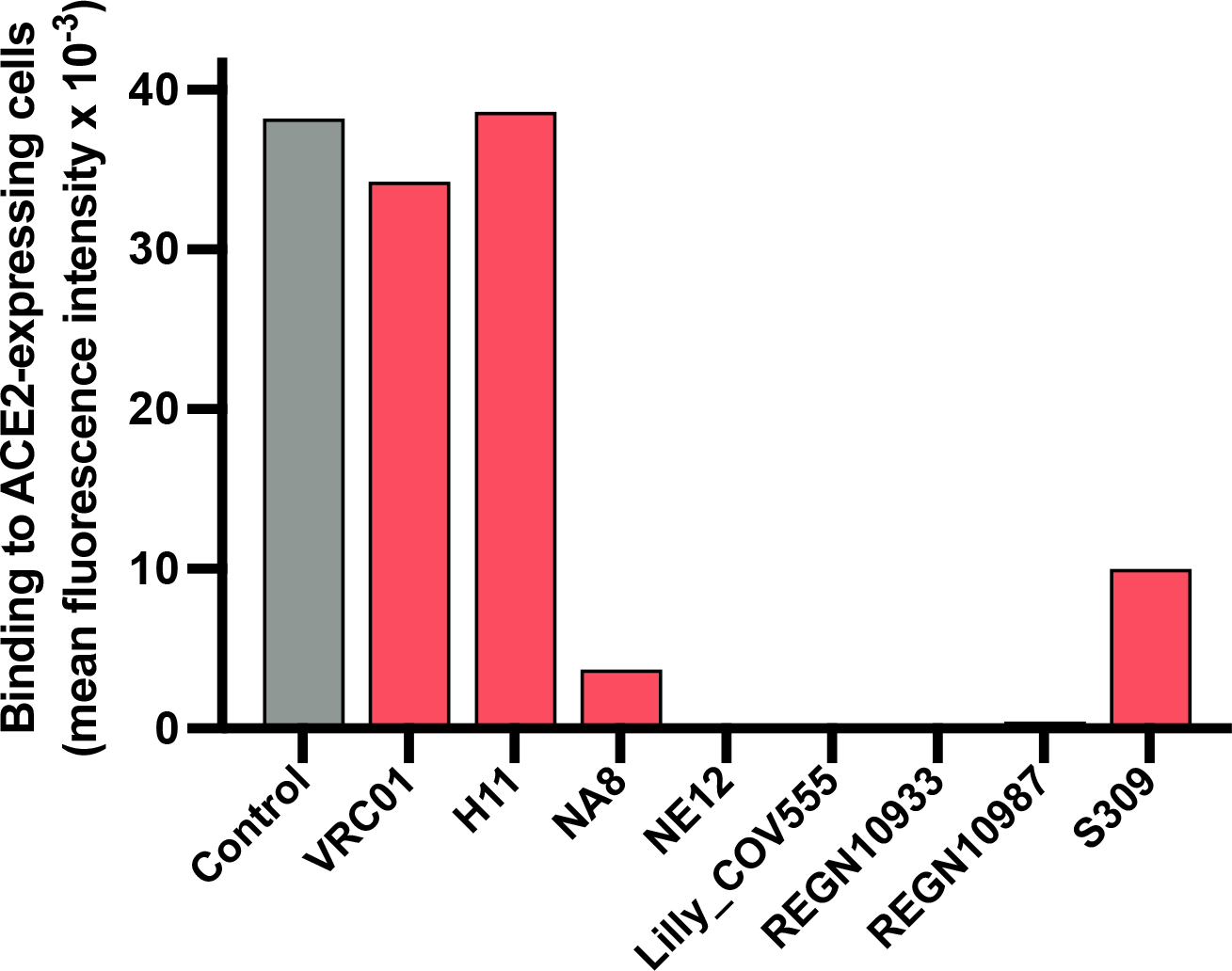
Competition of various mAbs with S-6P trimer binding to ACE-2-positive cells. Ability of monoclonal antibodies against the SARS-CoV-2 spike to block the interaction of the S-6P trimer to ACE2-expressing HEK293 cells. The trimer was directly labeled with phycoerythrin (PE) and pre-incubated with each of the mAbs or with PBS buffer (control) for 30 min before incubation with the cells for 30 min.

**Extended Data Figure 15.**
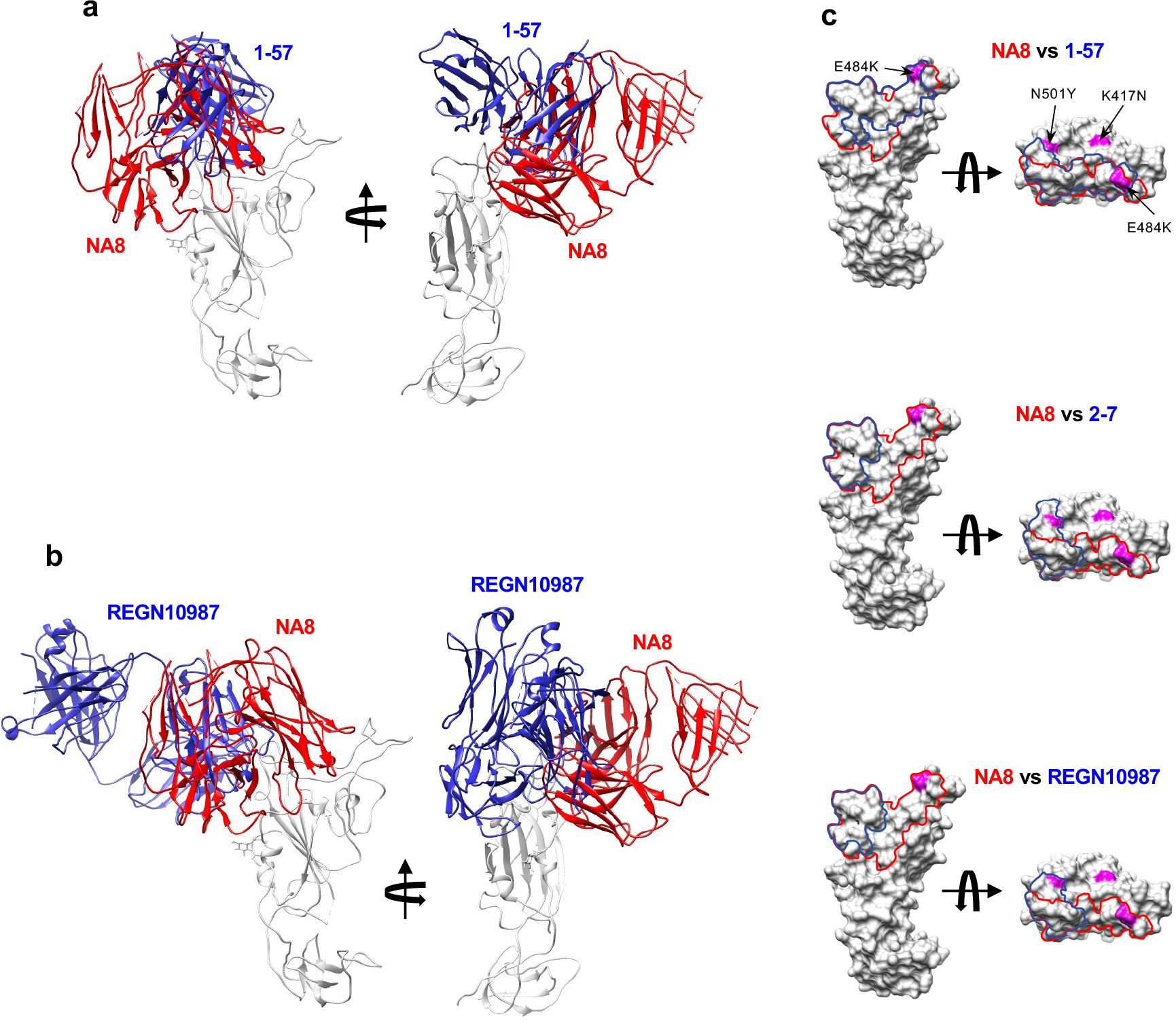
**Comparison of binding modes and epitopes of mAb NA8 and selected RBD-directed antibodies capable of neutralizing the B.1.351 variant.**^6^ **a**, Cartoon representations of aligned RBD-Fab complexes for NA8 and mAb 1-57 (from PDB 7l39). **b**, Cartoon representations of aligned RBD-Fab complexes for NA8 and mAb REGN10987 (from PDB 6xdg). **c**, The RBD is shown in surface representation with the epitopes of NA8 and mAbs 1-57, 2-7 and REGN10987 outlined with red and blue curves, respectively. Surface areas corresponding to the RBD residues mutated in the B.1.351 variant are colored in magenta.

**Extended Data Figure 16.**
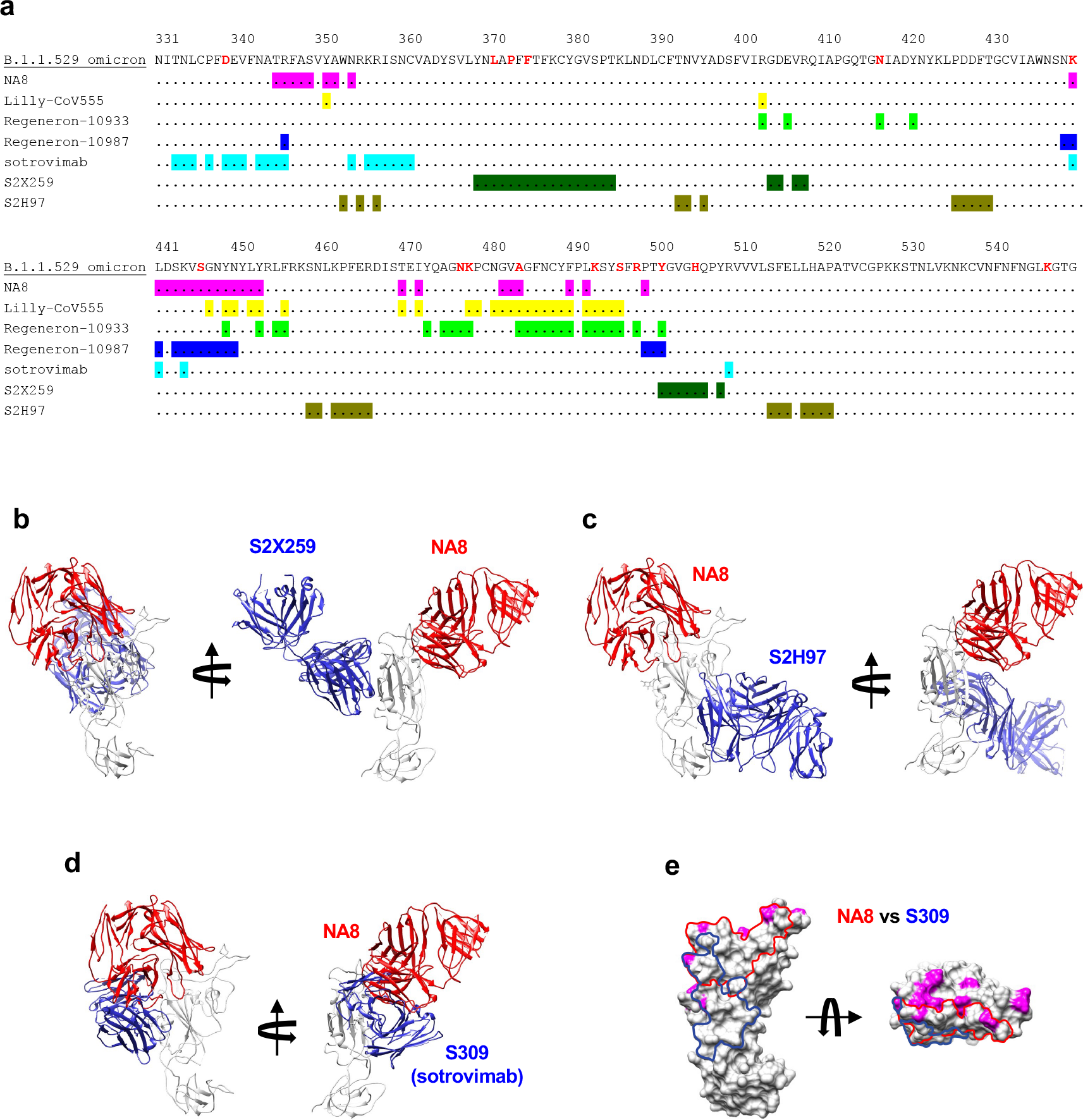
Comparison of epitopes and binding modes of mAb NA8 and known RBD-directed antibodies capable of neutralizing the B.1.1.529.1/Omicron variant. **a**, Sequence of RBD with B.1.1.529.1 mutations (colored red) with highlighted epitopes of mAbs. **b-d**, Cartoon representations of aligned RBD-Fab complexes for NA8 and mAb S2X259 (from PDB 7m7w) (a), mAb S2H97 (from PDB 7m7w) (b) and mAb S309 (sotrovimab) (from PDB 6wps). **e**, The RBD is shown in surface representation with the epitopes of NA8 and S309 outlined with red and blue curves, respectively. Surface areas corresponding to the RBD residues mutated in the B.1.1.529.1/Omicron variant are colored in magenta.

**Extended Data Table 1.**
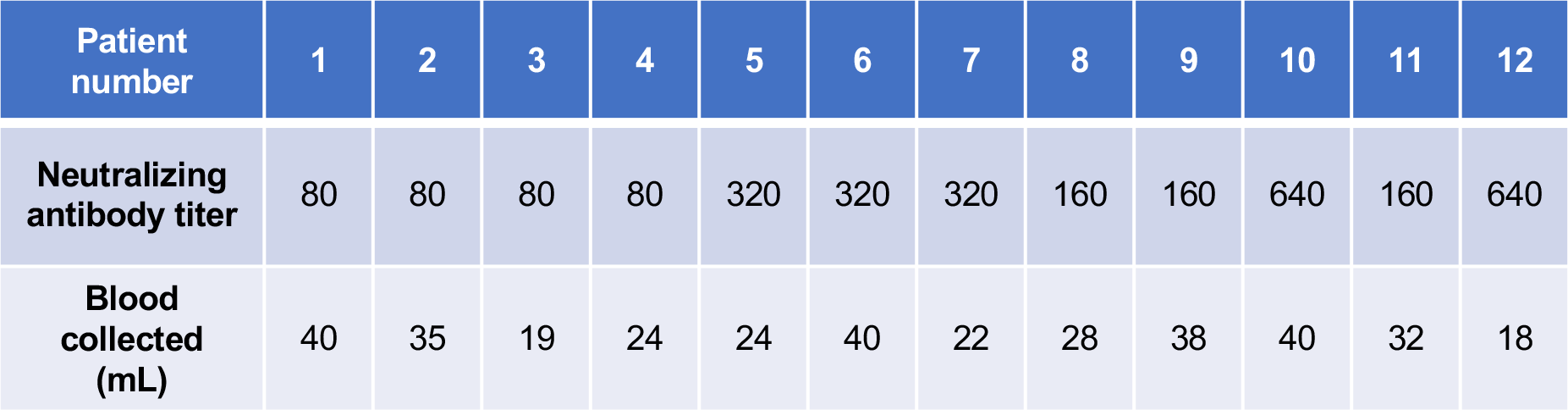
Neutralizing antibody titers in 12 COVID-19 convalescent plasma donors.

| Patient number | 1 | 2 | 3 | 4 | 5 | 6 | 7 | 8 | 9 | 10 | 11 | 12 |
| --- | --- | --- | --- | --- | --- | --- | --- | --- | --- | --- | --- | --- |
| Neutralizing antibody titer | 80 | 80 | 80 | 80 | 320 | 320 | 320 | 160 | 160 | 640 | 160 | 640 |
| Blood collected (mL) | 40 | 35 | 19 | 24 | 24 | 40 | 22 | 28 | 38 | 40 | 32 | 18 |

**Extended Data Table 2.** Binding affinity of anti-SARS-CoV-2 Fabs to the S-2P trimer, as measured by surface plasmon resonance.

| **Fab** | **$K_D$ (M)** | **$k_d$ (s<sup>-1</sup>)** | **$k_a$ (1/Ms)** |
| --- | --- | --- | --- |
| **1G8** | 6.2e-10 | 2.0e-4 | 4.6e5 |
| **3F6** | 4.3e-10 | 8.2e-4 | 3.9e6 |
| **4B6** | 8.7e-10 | 1.1e-3 | 1.3e6 |
| **D12** | 7.2e-10 | 8.7e-4 | 1.3e6 |
| **NA8** | 4.6 e-10 | 1.2e-3 | 2.8e6 |
| **2A12** | 3.0e-10 | 8.0e-4 | 2.7e6 |
| **NE8** | 1.8e-10 | 5.7e-3 | 3.1e6 |
| **NE12** | 6.1e-10* | 6.5e-4 | 1.3e6 |
| **NG3** | 2.2e-10 | 2.0e-4 | 1.0e6 |
| **4B7** | 1.1e-9 | 1.7e-3 | 1.6e6 |
| **A7** | 3.9e-10 | 7.8e-4 | 2.2e6 |
| **H11** | 6.5e-9 | 1.7e-3 | 2.7e5 |

\*Constants were determined using BiaEvaluation software. Constants are the average of two different analyses. Only one reliable data set was obtained for 4B7.

**Extended Data Table 3.**
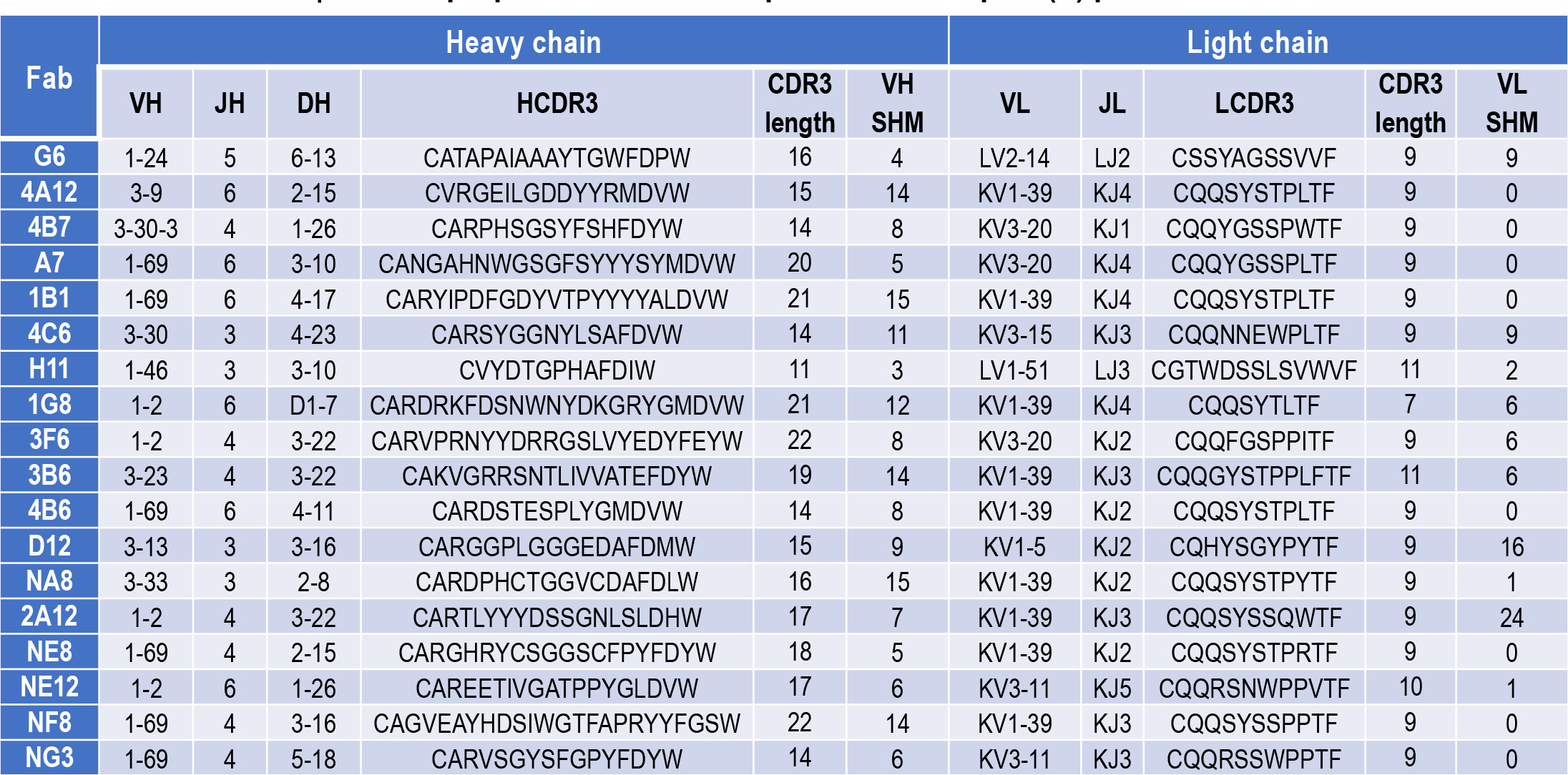
Genetic properties of 18 Fabs specific for the spike (S) protein of SARS-CoV-2

**Extended Data Table 4.** Cryo-EM data collection and analysis statistics.

| S-6P in complex<br>with Fab NE12 |  |  | S-6P in complex<br>with Fab NA8 |  |
| --- | --- | --- | --- | --- |
| Data collection and processing |  |  |  |  |
| Magnification | 105,000 |  | 105,000 |  |
| Voltage (kV) | 300 |  | 300 |  |
| Electron exposure (e<sup>-</sup>/Å<sup>2</sup>) | 40 |  | 40 |  |
| Defocus range (μm) | -1.0 to -2.5 |  | -1.0 to -2.5 |  |
| Pixel size (Å) | 0.873 |  | 0.873 |  |
| Symmetry imposed | C1 |  | C1 |  |
| Initial particle images (no.) | 2,253,882 |  | 2,870,476 |  |
| Final particle images (no.) | 201,607 |  | 454,910 |  |
|  | Consensus map<br>EMDB-xxxx | Local map<br>EMDB-xxxx<br>PDB xxxx | Consensus map<br>EMDB-xxxx | Local map<br>EMDB-xxxx<br>PDB xxxx |
| Map resolution (Å) | 3.1 | 3.4 | 2.9 | 3.5 |
| FSC threshold | 0.143 |  | 0.143 |  |
| Map resolution range (Å) | 1.9-6.8 | 3.1-6.8 | 1.9-8.0 | 3.1-6.8 |
| Refinement |  |  |  |  |
| Initial model used (PDB code) |  | 6xlu |  | 6xlu |
| Model resolution (Å) |  | 3.4 |  | 3.5 |
| FSC threshold |  | 0.5 |  | 0.5 |
| Map sharpening *B* factor (Å<sup>2</sup>) | -44.4 | -125.7 | -53.3 | -165.1 |
| Model composition |  |  |  |  |
| Non-hydrogen atoms |  | 6563 |  | 6185 |
| Protein residues |  | 829 |  | 779 |
| Ligands (glycans) |  | 3 |  | 4 |
| Water |  | 0 |  | 0 |
| *B* factors (Å<sup>2</sup>) (mean) |  |  |  |  |
| Protein |  | 58.9 |  | 64.2 |
| Ligand |  | 82.1 |  | 96.6 |
| Water |  | N/A |  | N/A |
| R.m.s. deviations |  |  |  |  |
| Bond lengths (Å) |  | 0.007 |  | 0.005 |
| Bond angles (°) |  | 0.926 |  | 0.68 |
| Validation |  |  |  |  |
| MolProbity score |  | 1.44 |  | 1.81 |
| Clash score |  | 1.55 |  | 6.52 |
| Poor rotamers (%) |  | 0.28 |  | 0 |
| Ramachandran plot |  |  |  |  |
| Favored (%) |  | 90.34 |  | 92.85 |
| Allowed (%) |  | 9.53 |  | 7.02 |
| Disallowed (%) |  | 0.13 |  | 0.13 |

